# Effectiveness of mRNA-1273 against SARS-CoV-2 omicron and delta variants

**DOI:** 10.1101/2022.01.07.22268919

**Authors:** Hung Fu Tseng, Bradley K. Ackerson, Yi Luo, Lina S. Sy, Carla A. Talarico, Yun Tian, Katia J. Bruxvoort, Julia E. Tubert, Ana Florea, Jennifer H. Ku, Gina S. Lee, Soon Kyu Choi, Harpreet S. Takhar, Michael Aragones, Lei Qian

**Author notes:** **Corresponding Author:** Hung Fu Tseng, Department of Research and Evaluation, Kaiser Permanente Southern California, 100 S Los Robles, Pasadena, CA 91101.

## Abstract

SARS-CoV-2 omicron (B.1.1.529) variant is highly transmissible with potential immune escape. We conducted a test-negative case-control study to evaluate mRNA-1273 vaccine effectiveness (VE) against infection and hospitalization with omicron or delta. The large, diverse study population included 26,683 SARS-CoV-2 test-positive cases with variant determined by spike gene status (16% delta, 84% omicron). The 2-dose VE against omicron infection at 14-90 days was 44.0% (95% CI, 35.1–51.6%) but declined quickly. The 3-dose VE was 93.7% (92.2–94.9%) and 86.0% (78.1–91.1%) against delta infection and 71.6% (69.7–73.4%) and 47.4% (40.5–53.5%) against omicron infection at 14-60 days and >60 days, respectively. The 3-dose VE was 29.4% (0.3–50.0%) against omicron infection in immunocompromised individuals. The 3-dose VE against hospitalization with delta or omicron was >99%. Our findings demonstrate high, durable 3-dose VE against delta infection but lower effectiveness against omicron infection, particularly among immunocompromised people. However, 3-dose VE was high against hospitalization with delta or omicron.

## Introduction

The recently emerged severe acute respiratory syndrome coronavirus 2 (SARS-CoV-2) omicron (B.1.1.529) variant contains multiple novel spike (S) protein mutations, raising concerns about escape from naturally acquired or vaccine-elicited immunity.^1^ Several *in vitro* studies reported reduced vaccine-induced neutralization activity against omicron.^2,3^ Specifically, sera from individuals vaccinated with 2 doses of mRNA coronavirus disease 2019 (COVID-19) vaccines, including mRNA-1273 (Moderna COVID-19 vaccine), showed substantial reductions in neutralization activity against omicron compared with wild-type SARS-CoV-2.^2,4,5^ However, an mRNA-1273 booster increased neutralization activity against omicron, albeit lower than wild-type.^2,3^ We previously reported high and durable vaccine effectiveness (VE) of mRNA-1273 against infection and hospitalization from COVID-19 caused by other emerging SARS-CoV-2 variants, including delta (B.1.617.2).^6^ While limited data are available on real-world VE of mRNA-1273 against omicron, an analysis of a US pharmacy-based testing program found that the likelihood of vaccination with 3 mRNA-1273 vaccine doses (vs unvaccinated) was significantly lower among omicron symptomatic infections (odds ratio, 0.31) than SARS-CoV-2-negative controls.^7^ Another US study during an omicron-predominant period found that receipt of a third mRNA vaccine dose was 90% effective in preventing COVID-19-associated hospitalization.^8^

As the omicron BA.1 sub-lineage has a deletion at positions 69-70, initial omicron-positive specimens exhibit S-gene target failure (SGTF). To provide timely results for these analyses, we used SGTF as a marker for omicron in specimens collected during December 2021. The US Food and Drug Administration (FDA) and World Health Organization advised that SGTF from select COVID-19 RT-PCR assays, including the Thermo Fisher TaqPath™ COVID-19 Combo kits, can be used as a screening method for omicron;^9,10^ SGTF has served as a proxy in the United Kingdom for identifying omicron.^11,12^ In Southern California, where delta was the dominant strain before omicron^13^ and the proportion of SGTF among SARS-CoV-2 positive specimens increased from 1.2% to 94.1% from 12/06/2021 to 12/31/2021, SGTF can be used as a proxy for omicron sub-lineage BA.1, while positive specimens negative for SGTF can be considered delta. Using electronic health records (EHR) from the Kaiser Permanente Southern California (KPSC) health care system in the United States, we conducted a test-negative case-control study to evaluate the VE of mRNA-1273 against infection and hospitalization with omicron and delta.

## Results

The study included 26,683 cases with SGTF status available. Based on whole genome sequencing results received for a subset of 1,383 positive specimens, we confirmed that all 704 cases exhibiting SGTF were omicron (100%), and 673 of the 679 SGTF negative cases were delta (99.1%), with a kappa 0.991. The sensitivity and specificity of SGTF in predicting omicron was 99.2% and 100%, respectively. Of the 26,683 cases, 11,483 (43.0%) individuals were unvaccinated (2,883 delta, 8,600 omicron), and 15,200 (57.0%) were vaccinated (1,431 delta, 13,769 omicron; 416 vaccinated with 1 dose, 12,029 vaccinated with 2 doses, 2,755 vaccinated with 3 doses). The flow chart depicting selection steps is provided (Fig. 1). The distribution of covariates by test outcomes, separated by variant type, is summarized in Table 1 (2-dose and 3-dose analyses) and Supplementary Table 1 (1-dose analysis).

**Fig. 1:**
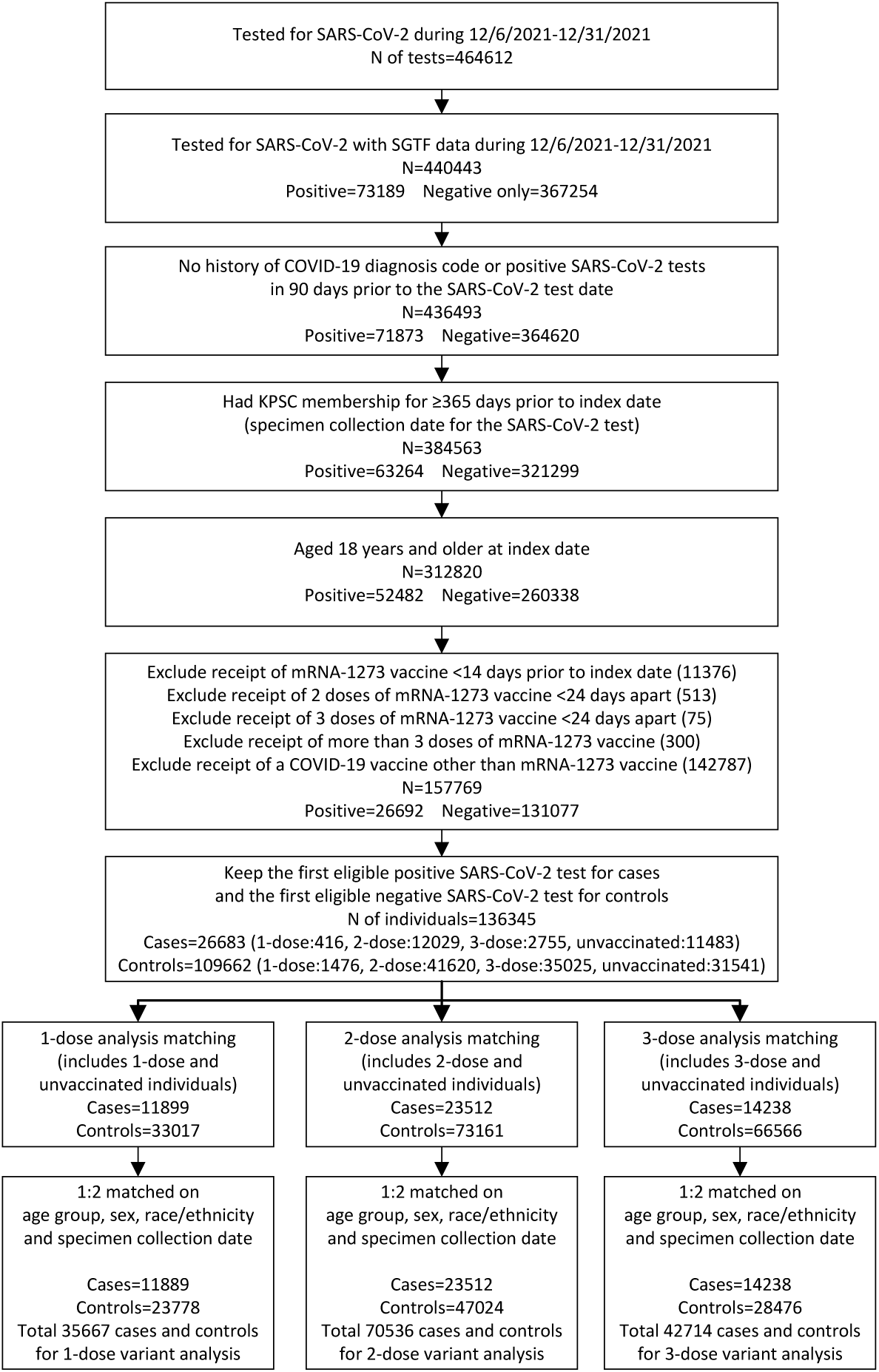
Flowchart of Selection of Cases and Controls The figure depicts the steps for selection of 26,683 cases and 109,662 controls by inclusion and exclusion criteria, and subsequent matching in 1-dose, 2-dose, and 3-dose analyses.

**Table 1.**
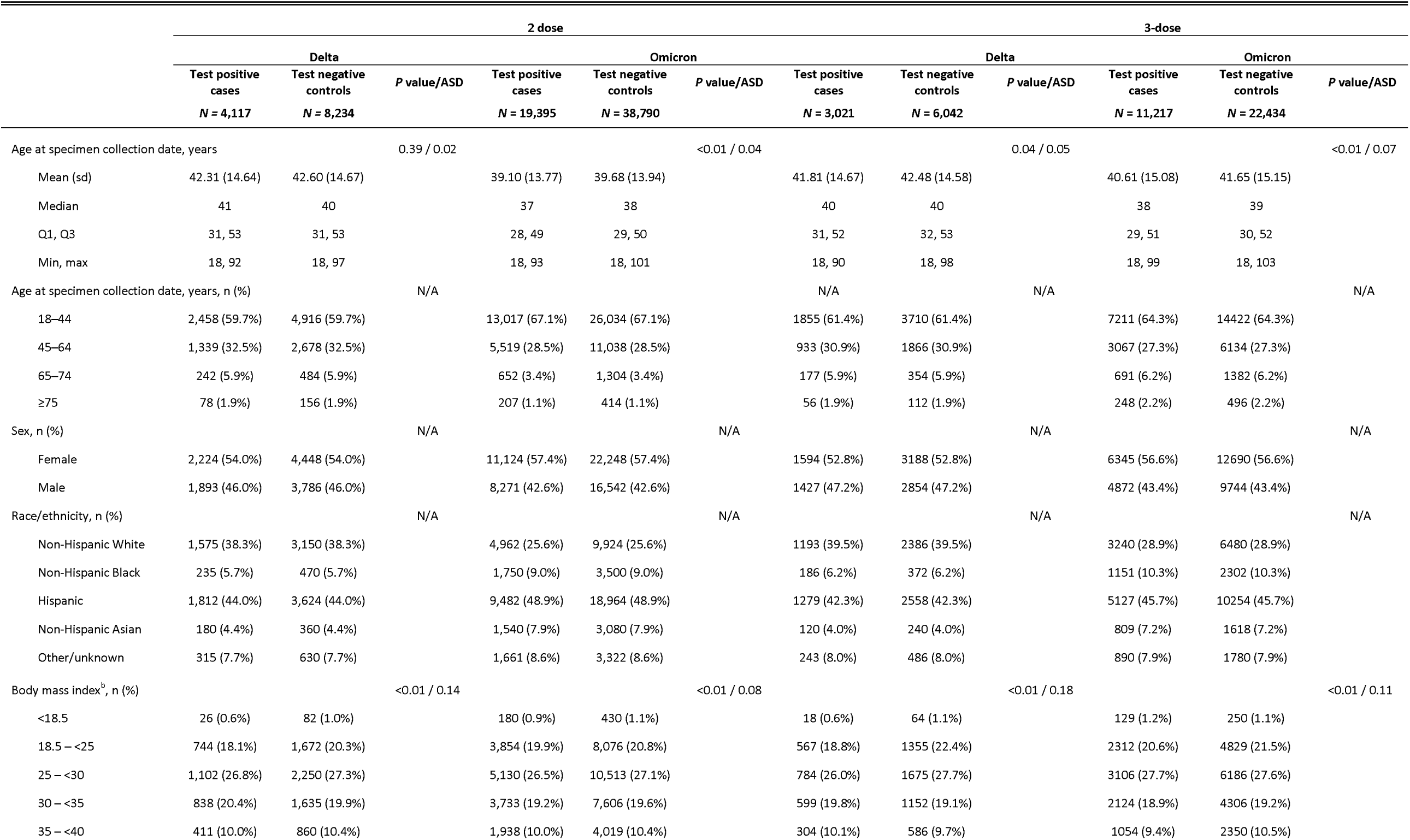
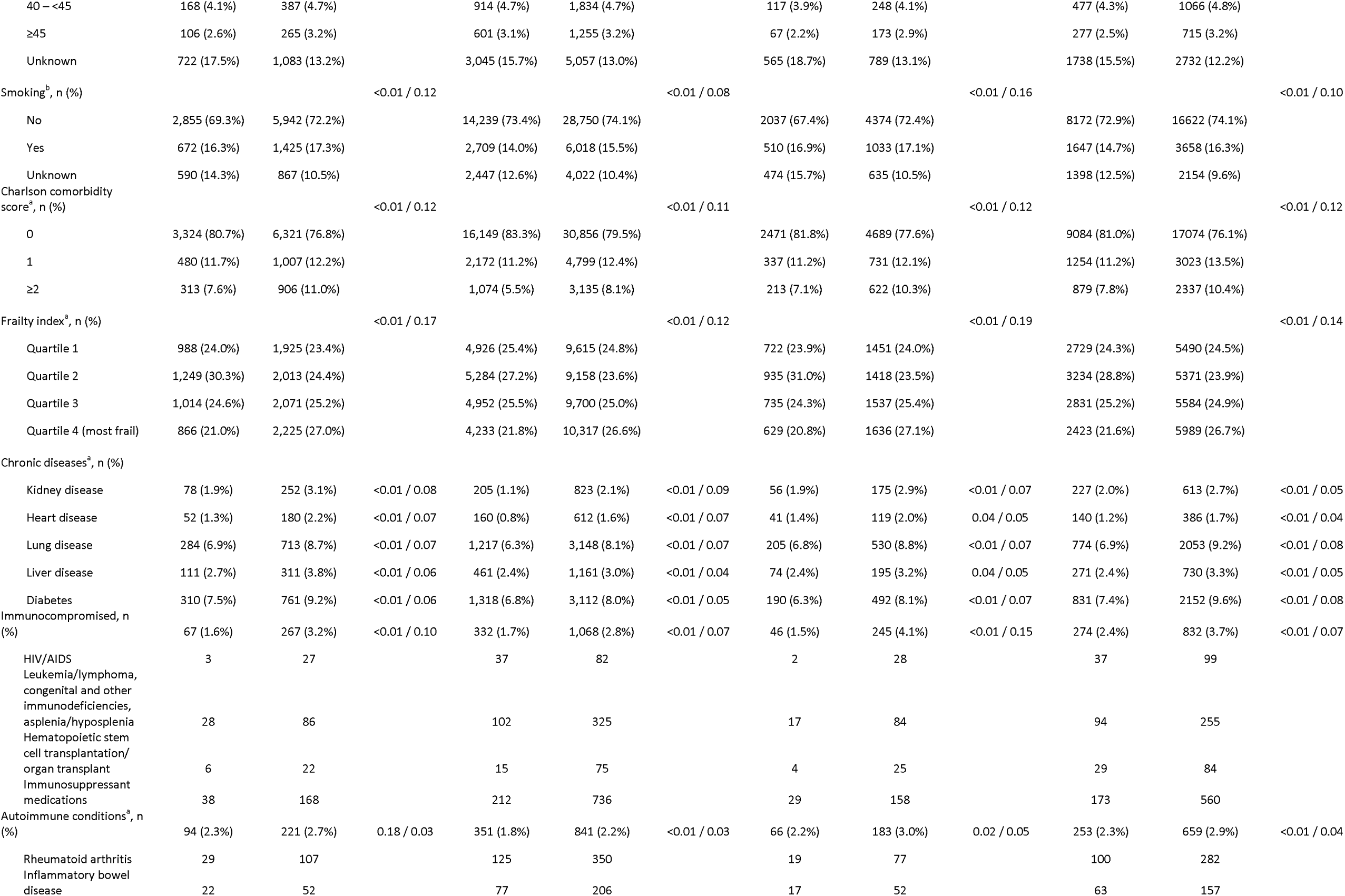
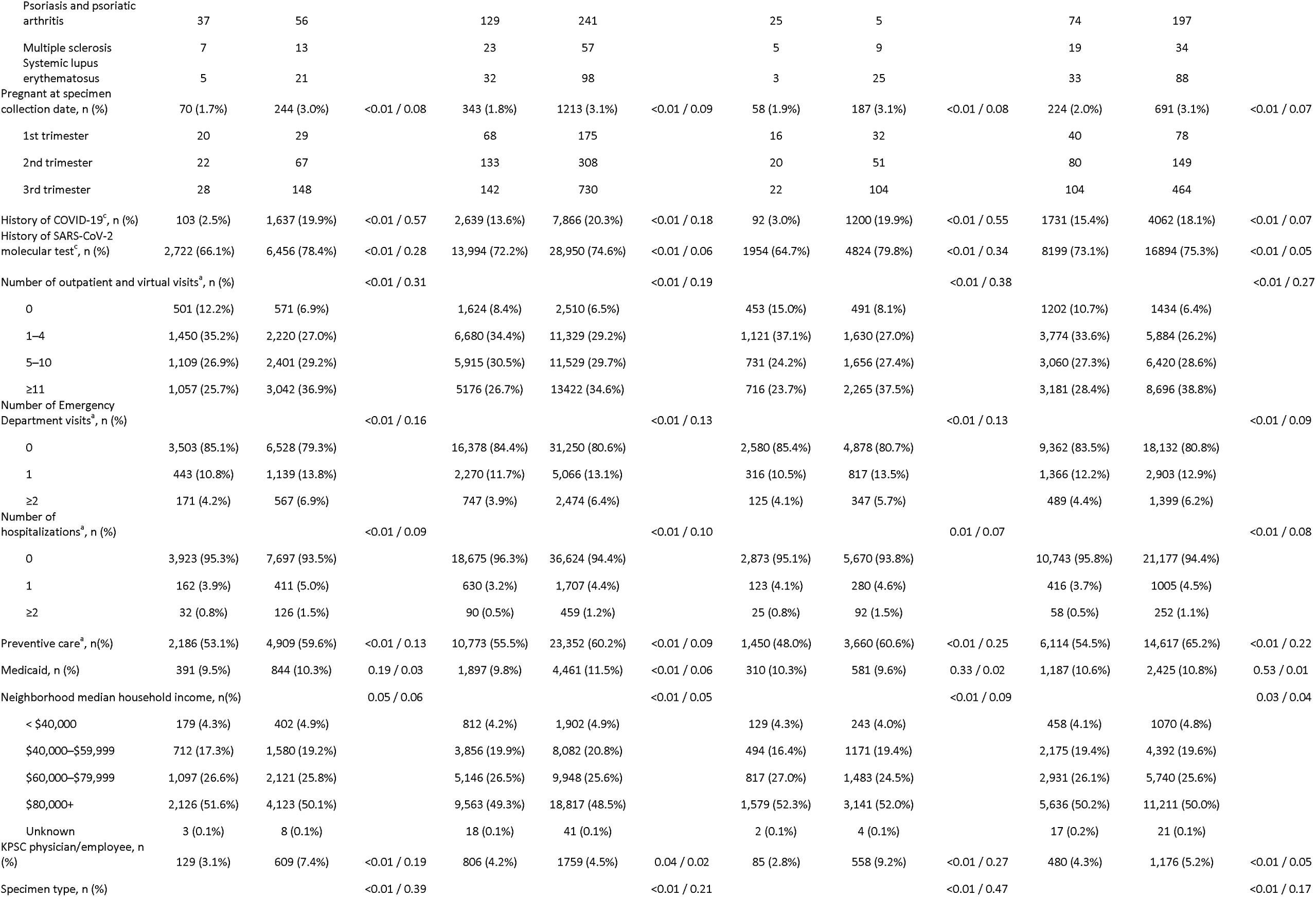
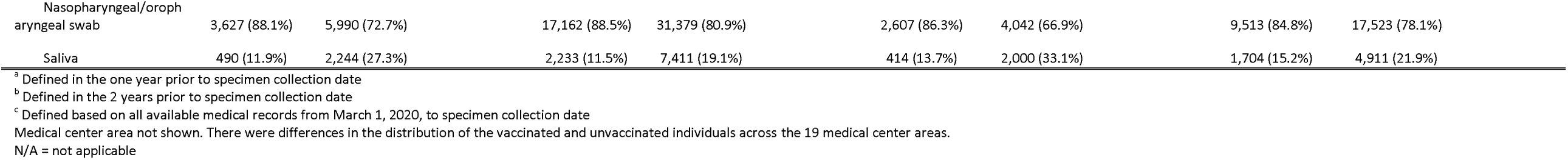
Characteristics of SARS-CoV-2 cases and controls, by variant

|  | 2 dose |  |  |  |  |  | 3-dose |  |  |  |  |  |
| --- | --- | --- | --- | --- | --- | --- | --- | --- | --- | --- | --- | --- |
|  | Delta |  |  | Omicron |  |  | Delta |  |  | Omicron |  |  |
|  | Test positive cases<br>*N* = 4,117 | Test negative controls<br>*N* = 8,234 | *P* value/ASD | Test positive cases<br>*N* = 19,395 | Test negative controls<br>*N* = 38,790 | *P* value/ASD | Test positive cases<br>*N* = 3,021 | Test negative controls<br>*N* = 6,042 | *P* value/ASD | Test positive cases<br>*N* = 11,217 | Test negative controls<br>*N* = 22,434 | *P* value/ASD |
| Age at specimen collection date, years |  |  | 0.39 / 0.02 |  |  | <0.01 / 0.04 |  |  | 0.04 / 0.05 |  |  | <0.01 / 0.07 |
| Mean (sd) | 42.31 (14.64) | 42.60 (14.67) |  | 39.10 (13.77) | 39.68 (13.94) |  | 41.81 (14.67) | 42.48 (14.58) |  | 40.61 (15.08) | 41.65 (15.15) |  |
| Median | 41 | 40 |  | 37 | 38 |  | 40 | 40 |  | 38 | 39 |  |
| Q1, Q3 | 31, 53 | 31, 53 |  | 28, 49 | 29, 50 |  | 31, 52 | 32, 53 |  | 29, 51 | 30, 52 |  |
| Min, max | 18, 92 | 18, 97 |  | 18, 93 | 18, 101 |  | 18, 90 | 18, 98 |  | 18, 99 | 18, 103 |  |
| Age at specimen collection date, years, n (%) |  |  | N/A |  |  | N/A |  |  | N/A |  |  | N/A |
| 18–44 | 2,458 (59.7%) | 4,916 (59.7%) |  | 13,017 (67.1%) | 26,034 (67.1%) |  | 1855 (61.4%) | 3710 (61.4%) |  | 7211 (64.3%) | 14422 (64.3%) |  |
| 45–64 | 1,339 (32.5%) | 2,678 (32.5%) |  | 5,519 (28.5%) | 11,038 (28.5%) |  | 933 (30.9%) | 1866 (30.9%) |  | 3067 (27.3%) | 6134 (27.3%) |  |
| 65–74 | 242 (5.9%) | 484 (5.9%) |  | 652 (3.4%) | 1,304 (3.4%) |  | 177 (5.9%) | 354 (5.9%) |  | 691 (6.2%) | 1382 (6.2%) |  |
| ≥75 | 78 (1.9%) | 156 (1.9%) |  | 207 (1.1%) | 414 (1.1%) |  | 56 (1.9%) | 112 (1.9%) |  | 248 (2.2%) | 496 (2.2%) |  |
| Sex, n (%) |  |  | N/A |  |  | N/A |  |  | N/A |  |  | N/A |
| Female | 2,224 (54.0%) | 4,448 (54.0%) |  | 11,124 (57.4%) | 22,248 (57.4%) |  | 1594 (52.8%) | 3188 (52.8%) |  | 6345 (56.6%) | 12690 (56.6%) |  |
| Male | 1,893 (46.0%) | 3,786 (46.0%) |  | 8,271 (42.6%) | 16,542 (42.6%) |  | 1427 (47.2%) | 2854 (47.2%) |  | 4872 (43.4%) | 9744 (43.4%) |  |
| Race/ethnicity, n (%) |  |  | N/A |  |  | N/A |  |  | N/A |  |  | N/A |
| Non-Hispanic White | 1,575 (38.3%) | 3,150 (38.3%) |  | 4,962 (25.6%) | 9,924 (25.6%) |  | 1193 (39.5%) | 2386 (39.5%) |  | 3240 (28.9%) | 6480 (28.9%) |  |
| Non-Hispanic Black | 235 (5.7%) | 470 (5.7%) |  | 1,750 (9.0%) | 3,500 (9.0%) |  | 186 (6.2%) | 372 (6.2%) |  | 1151 (10.3%) | 2302 (10.3%) |  |
| Hispanic | 1,812 (44.0%) | 3,624 (44.0%) |  | 9,482 (48.9%) | 18,964 (48.9%) |  | 1279 (42.3%) | 2558 (42.3%) |  | 5127 (45.7%) | 10254 (45.7%) |  |
| Non-Hispanic Asian | 180 (4.4%) | 360 (4.4%) |  | 1,540 (7.9%) | 3,080 (7.9%) |  | 120 (4.0%) | 240 (4.0%) |  | 809 (7.2%) | 1618 (7.2%) |  |
| Other/unknown | 315 (7.7%) | 630 (7.7%) |  | 1,661 (8.6%) | 3,322 (8.6%) |  | 243 (8.0%) | 486 (8.0%) |  | 890 (7.9%) | 1780 (7.9%) |  |
| Body mass index<sup>b</sup>, n (%) |  |  | <0.01 / 0.14 |  |  | <0.01 / 0.08 |  |  | <0.01 / 0.18 |  |  | <0.01 / 0.11 |
| <18.5 | 26 (0.6%) | 82 (1.0%) |  | 180 (0.9%) | 430 (1.1%) |  | 18 (0.6%) | 64 (1.1%) |  | 129 (1.2%) | 250 (1.1%) |  |
| 18.5 – <25 | 744 (18.1%) | 1,672 (20.3%) |  | 3,854 (19.9%) | 8,076 (20.8%) |  | 567 (18.8%) | 1355 (22.4%) |  | 2312 (20.6%) | 4829 (21.5%) |  |
| 25 – <30 | 1,102 (26.8%) | 2,250 (27.3%) |  | 5,130 (26.5%) | 10,513 (27.1%) |  | 784 (26.0%) | 1675 (27.7%) |  | 3106 (27.7%) | 6186 (27.6%) |  |
| 30 – <35 | 838 (20.4%) | 1,635 (19.9%) |  | 3,733 (19.2%) | 7,606 (19.6%) |  | 599 (19.8%) | 1152 (19.1%) |  | 2124 (18.9%) | 4306 (19.2%) |  |
| 35 – <40 | 411 (10.0%) | 860 (10.4%) |  | 1,938 (10.0%) | 4,019 (10.4%) |  | 304 (10.1%) | 586 (9.7%) |  | 1054 (9.4%) | 2350 (10.5%) |  |
| 40 – <45 | 168 (4.1%) | 387 (4.7%) |  | 914 (4.7%) | 1,834 (4.7%) |  | 117 (3.9%) | 248 (4.1%) |  | 477 (4.3%) | 1066 (4.8%) |  |
| ≥45 | 106 (2.6%) | 265 (3.2%) |  | 601 (3.1%) | 1,255 (3.2%) |  | 67 (2.2%) | 173 (2.9%) |  | 277 (2.5%) | 715 (3.2%) |  |
| Unknown | 722 (17.5%) | 1,083 (13.2%) |  | 3,045 (15.7%) | 5,057 (13.0%) |  | 565 (18.7%) | 789 (13.1%) |  | 1738 (15.5%) | 2732 (12.2%) |  |
| Smoking<sup>b</sup>, n (%) |  |  | <0.01 / 0.12 |  |  | <0.01 / 0.08 |  |  | <0.01 / 0.16 |  |  | <0.01 / 0.10 |
| No | 2,855 (69.3%) | 5,942 (72.2%) |  | 14,239 (73.4%) | 28,750 (74.1%) |  | 2037 (67.4%) | 4374 (72.4%) |  | 8172 (72.9%) | 16622 (74.1%) |  |
| Yes | 672 (16.3%) | 1,425 (17.3%) |  | 2,709 (14.0%) | 6,018 (15.5%) |  | 510 (16.9%) | 1033 (17.1%) |  | 1647 (14.7%) | 3658 (16.3%) |  |
| Unknown | 590 (14.3%) | 867 (10.5%) |  | 2,447 (12.6%) | 4,022 (10.4%) |  | 474 (15.7%) | 635 (10.5%) |  | 1398 (12.5%) | 2154 (9.6%) |  |
| Charlson comorbidity score<sup>a</sup>, n (%) |  |  | <0.01 / 0.12 |  |  | <0.01 / 0.11 |  |  | <0.01 / 0.12 |  |  | <0.01 / 0.12 |
| 0 | 3,324 (80.7%) | 6,321 (76.8%) |  | 16,149 (83.3%) | 30,856 (79.5%) |  | 2471 (81.8%) | 4689 (77.6%) |  | 9084 (81.0%) | 17074 (76.1%) |  |
| 1 | 480 (11.7%) | 1,007 (12.2%) |  | 2,172 (11.2%) | 4,799 (12.4%) |  | 337 (11.2%) | 731 (12.1%) |  | 1254 (11.2%) | 3023 (13.5%) |  |
| ≥2 | 313 (7.6%) | 906 (11.0%) |  | 1,074 (5.5%) | 3,135 (8.1%) |  | 213 (7.1%) | 622 (10.3%) |  | 879 (7.8%) | 2337 (10.4%) |  |
| Frailty index<sup>a</sup>, n (%) |  |  | <0.01 / 0.17 |  |  | <0.01 / 0.12 |  |  | <0.01 / 0.19 |  |  | <0.01 / 0.14 |
| Quartile 1 | 988 (24.0%) | 1,925 (23.4%) |  | 4,926 (25.4%) | 9,615 (24.8%) |  | 722 (23.9%) | 1451 (24.0%) |  | 2729 (24.3%) | 5490 (24.5%) |  |
| Quartile 2 | 1,249 (30.3%) | 2,013 (24.4%) |  | 5,284 (27.2%) | 9,158 (23.6%) |  | 935 (31.0%) | 1418 (23.5%) |  | 3234 (28.8%) | 5371 (23.9%) |  |
| Quartile 3 | 1,014 (24.6%) | 2,071 (25.2%) |  | 4,952 (25.5%) | 9,700 (25.0%) |  | 735 (24.3%) | 1537 (25.4%) |  | 2831 (25.2%) | 5584 (24.9%) |  |
| Quartile 4 (most frail) | 866 (21.0%) | 2,225 (27.0%) |  | 4,233 (21.8%) | 10,317 (26.6%) |  | 629 (20.8%) | 1636 (27.1%) |  | 2423 (21.6%) | 5989 (26.7%) |  |
| Chronic diseases<sup>a</sup>, n (%) |  |  |  |  |  |  |  |  |  |  |  |  |
| Kidney disease | 78 (1.9%) | 252 (3.1%) | <0.01 / 0.08 | 205 (1.1%) | 823 (2.1%) | <0.01 / 0.09 | 56 (1.9%) | 175 (2.9%) | <0.01 / 0.07 | 227 (2.0%) | 613 (2.7%) | <0.01 / 0.05 |
| Heart disease | 52 (1.3%) | 180 (2.2%) | <0.01 / 0.07 | 160 (0.8%) | 612 (1.6%) | <0.01 / 0.07 | 41 (1.4%) | 119 (2.0%) | 0.04 / 0.05 | 140 (1.2%) | 386 (1.7%) | <0.01 / 0.04 |
| Lung disease | 284 (6.9%) | 713 (8.7%) | <0.01 / 0.07 | 1,217 (6.3%) | 3,148 (8.1%) | <0.01 / 0.07 | 205 (6.8%) | 530 (8.8%) | <0.01 / 0.07 | 774 (6.9%) | 2053 (9.2%) | <0.01 / 0.08 |
| Liver disease | 111 (2.7%) | 311 (3.8%) | <0.01 / 0.06 | 461 (2.4%) | 1,161 (3.0%) | <0.01 / 0.04 | 74 (2.4%) | 195 (3.2%) | 0.04 / 0.05 | 271 (2.4%) | 730 (3.3%) | <0.01 / 0.05 |
| Diabetes | 310 (7.5%) | 761 (9.2%) | <0.01 / 0.06 | 1,318 (6.8%) | 3,112 (8.0%) | <0.01 / 0.05 | 190 (6.3%) | 492 (8.1%) | <0.01 / 0.07 | 831 (7.4%) | 2152 (9.6%) | <0.01 / 0.08 |
| Immunocompromised, n (%) | 67 (1.6%) | 267 (3.2%) | <0.01 / 0.10 | 332 (1.7%) | 1,068 (2.8%) | <0.01 / 0.07 | 46 (1.5%) | 245 (4.1%) | <0.01 / 0.15 | 274 (2.4%) | 832 (3.7%) | <0.01 / 0.07 |
| HIV/AIDS | 3 | 27 |  | 37 | 82 |  | 2 | 28 |  | 37 | 99 |  |
| Leukemia/lymphoma, congenital and other immunodeficiencies, asplenia/hyposplenia | 28 | 86 |  | 102 | 325 |  | 17 | 84 |  | 94 | 255 |  |
| Hematopoietic stem cell transplantation/organ transplant | 6 | 22 |  | 15 | 75 |  | 4 | 25 |  | 29 | 84 |  |
| Immunosuppressant medications | 38 | 168 |  | 212 | 736 |  | 29 | 158 |  | 173 | 560 |  |
| Autoimmune conditions<sup>a</sup>, n (%) | 94 (2.3%) | 221 (2.7%) | 0.18 / 0.03 | 351 (1.8%) | 841 (2.2%) | <0.01 / 0.03 | 66 (2.2%) | 183 (3.0%) | 0.02 / 0.05 | 253 (2.3%) | 659 (2.9%) | <0.01 / 0.04 |
| Rheumatoid arthritis | 29 | 107 |  | 125 | 350 |  | 19 | 77 |  | 100 | 282 |  |
| Inflammatory bowel disease | 22 | 52 |  | 77 | 206 |  | 17 | 52 |  | 63 | 157 |  |
| Psoriasis and psoriatic arthritis | 37 | 56 |  | 129 | 241 |  | 25 | 5 |  | 74 | 197 |  |
| Multiple sclerosis | 7 | 13 |  | 23 | 57 |  | 5 | 9 |  | 19 | 34 |  |
| Systemic lupus erythematosus | 5 | 21 |  | 32 | 98 |  | 3 | 25 |  | 33 | 88 |  |
| Pregnant at specimen collection date, n (%) | 70 (1.7%) | 244 (3.0%) | <0.01 / 0.08 | 343 (1.8%) | 1213 (3.1%) | <0.01 / 0.09 | 58 (1.9%) | 187 (3.1%) | <0.01 / 0.08 | 224 (2.0%) | 691 (3.1%) | <0.01 / 0.07 |
| 1st trimester | 20 | 29 |  | 68 | 175 |  | 16 | 32 |  | 40 | 78 |  |
| 2nd trimester | 22 | 67 |  | 133 | 308 |  | 20 | 51 |  | 80 | 149 |  |
| 3rd trimester | 28 | 148 |  | 142 | 730 |  | 22 | 104 |  | 104 | 464 |  |
| History of COVID-19<sup>c</sup>, n (%) | 103 (2.5%) | 1,637 (19.9%) | <0.01 / 0.57 | 2,639 (13.6%) | 7,866 (20.3%) | <0.01 / 0.18 | 92 (3.0%) | 1200 (19.9%) | <0.01 / 0.55 | 1731 (15.4%) | 4062 (18.1%) | <0.01 / 0.07 |
| History of SARS-CoV-2 molecular test<sup>c</sup>, n (%) | 2,722 (66.1%) | 6,456 (78.4%) | <0.01 / 0.28 | 13,994 (72.2%) | 28,950 (74.6%) | <0.01 / 0.06 | 1954 (64.7%) | 4824 (79.8%) | <0.01 / 0.34 | 8199 (73.1%) | 16894 (75.3%) | <0.01 / 0.05 |
| Number of outpatient and virtual visits<sup>a</sup>, n (%) |  |  | <0.01 / 0.31 |  |  | <0.01 / 0.19 |  |  | <0.01 / 0.38 |  |  | <0.01 / 0.27 |
| 0 | 501 (12.2%) | 571 (6.9%) |  | 1,624 (8.4%) | 2,510 (6.5%) |  | 453 (15.0%) | 491 (8.1%) |  | 1202 (10.7%) | 1434 (6.4%) |  |
| 1–4 | 1,450 (35.2%) | 2,220 (27.0%) |  | 6,680 (34.4%) | 11,329 (29.2%) |  | 1,121 (37.1%) | 1,630 (27.0%) |  | 3,774 (33.6%) | 5,884 (26.2%) |  |
| 5–10 | 1,109 (26.9%) | 2,401 (29.2%) |  | 5,915 (30.5%) | 11,529 (29.7%) |  | 731 (24.2%) | 1,656 (27.4%) |  | 3,060 (27.3%) | 6,420 (28.6%) |  |
| ≥11 | 1,057 (25.7%) | 3,042 (36.9%) |  | 5176 (26.7%) | 13422 (34.6%) |  | 716 (23.7%) | 2,265 (37.5%) |  | 3,181 (28.4%) | 8,696 (38.8%) |  |
| Number of Emergency Department visits<sup>a</sup>, n (%) |  |  | <0.01 / 0.16 |  |  | <0.01 / 0.13 |  |  | <0.01 / 0.13 |  |  | <0.01 / 0.09 |
| 0 | 3,503 (85.1%) | 6,528 (79.3%) |  | 16,378 (84.4%) | 31,250 (80.6%) |  | 2,580 (85.4%) | 4,878 (80.7%) |  | 9,362 (83.5%) | 18,132 (80.8%) |  |
| 1 | 443 (10.8%) | 1,139 (13.8%) |  | 2,270 (11.7%) | 5,066 (13.1%) |  | 316 (10.5%) | 817 (13.5%) |  | 1,366 (12.2%) | 2,903 (12.9%) |  |
| ≥2 | 171 (4.2%) | 567 (6.9%) |  | 747 (3.9%) | 2,474 (6.4%) |  | 125 (4.1%) | 347 (5.7%) |  | 489 (4.4%) | 1,399 (6.2%) |  |
| Number of hospitalizations<sup>a</sup>, n (%) |  |  | <0.01 / 0.09 |  |  | <0.01 / 0.10 |  |  | 0.01 / 0.07 |  |  | <0.01 / 0.08 |
| 0 | 3,923 (95.3%) | 7,697 (93.5%) |  | 18,675 (96.3%) | 36,624 (94.4%) |  | 2,873 (95.1%) | 5,670 (93.8%) |  | 10,743 (95.8%) | 21,177 (94.4%) |  |
| 1 | 162 (3.9%) | 411 (5.0%) |  | 630 (3.2%) | 1,707 (4.4%) |  | 123 (4.1%) | 280 (4.6%) |  | 416 (3.7%) | 1005 (4.5%) |  |
| ≥2 | 32 (0.8%) | 126 (1.5%) |  | 90 (0.5%) | 459 (1.2%) |  | 25 (0.8%) | 92 (1.5%) |  | 58 (0.5%) | 252 (1.1%) |  |
| Preventive care<sup>a</sup>, n(%) | 2,186 (53.1%) | 4,909 (59.6%) | <0.01 / 0.13 | 10,773 (55.5%) | 23,352 (60.2%) | <0.01 / 0.09 | 1,450 (48.0%) | 3,660 (60.6%) | <0.01 / 0.25 | 6,114 (54.5%) | 14,617 (65.2%) | <0.01 / 0.22 |
| Medicaid, n (%) | 391 (9.5%) | 844 (10.3%) | 0.19 / 0.03 | 1,897 (9.8%) | 4,461 (11.5%) | <0.01 / 0.06 | 310 (10.3%) | 581 (9.6%) | 0.33 / 0.02 | 1,187 (10.6%) | 2,425 (10.8%) | 0.53 / 0.01 |
| Neighborhood median household income, n(%) |  |  | 0.05 / 0.06 |  |  | <0.01 / 0.05 |  |  | <0.01 / 0.09 |  |  | 0.03 / 0.04 |
| < \$40,000 | 179 (4.3%) | 402 (4.9%) | | 812 (4.2%) | 1,902 (4.9%) | | 129 (4.3%) | 243 (4.0%) | | 458 (4.1%) | 1070 (4.8%) | |
| \$40,000–\$59,999 | 712 (17.3%) | 1,580 (19.2%) | | 3,856 (19.9%) | 8,082 (20.8%) | | 494 (16.4%) | 1171 (19.4%) | | 2,175 (19.4%) | 4,392 (19.6%) | |
| \$60,000–\$79,999 | 1,097 (26.6%) | 2,121 (25.8%) | | 5,146 (26.5%) | 9,948 (25.6%) | | 817 (27.0%) | 1,483 (24.5%) | | 2,931 (26.1%) | 5,740 (25.6%) | |
| \$80,000+ | 2,126 (51.6%) | 4,123 (50.1%) | | 9,563 (49.3%) | 18,817 (48.5%) | | 1,579 (52.3%) | 3,141 (52.0%) | | 5,636 (50.2%) | 11,211 (50.0%) | |
| Unknown | 3 (0.1%) | 8 (0.1%) |  | 18 (0.1%) | 41 (0.1%) |  | 2 (0.1%) | 4 (0.1%) |  | 17 (0.2%) | 21 (0.1%) |  |
| KPSC physician/employee, n (%) | 129 (3.1%) | 609 (7.4%) | <0.01 / 0.19 | 806 (4.2%) | 1759 (4.5%) | 0.04 / 0.02 | 85 (2.8%) | 558 (9.2%) | <0.01 / 0.27 | 480 (4.3%) | 1,176 (5.2%) | <0.01 / 0.05 |
| Specimen type, n (%) |  |  | <0.01 / 0.39 |  |  | <0.01 / 0.21 |  |  | <0.01 / 0.47 |  |  | <0.01 / 0.17 |

|  |  |  |  |  |  |  |  |  |
| --- | --- | --- | --- | --- | --- | --- | --- | --- |
| Nasopharyngeal/oropharyngeal swab | 3,627 (88.1%) | 5,990 (72.7%) | 17,162 (88.5%) | 31,379 (80.9%) | 2,607 (86.3%) | 4,042 (66.9%) | 9,513 (84.8%) | 17,523 (78.1%) |
| Saliva | 490 (11.9%) | 2,244 (27.3%) | 2,233 (11.5%) | 7,411 (19.1%) | 414 (13.7%) | 2,000 (33.1%) | 1,704 (15.2%) | 4,911 (21.9%) |

<sup>a</sup> Defined in the one year prior to specimen collection date
<sup>b</sup> Defined in the 2 years prior to specimen collection date
<sup>c</sup> Defined based on all available medical records from March 1, 2020, to specimen collection date
Medical center area not shown. There were differences in the distribution of the vaccinated and unvaccinated individuals across the 19 medical center areas.
N/A = not applicable

Omicron cases more frequently had a history of COVID-19 than delta cases. In the 2-dose and 3-dose analyses, 13.6% and 15.4% of omicron cases in the 2-dose and 3-dose analyses, respectively, had a history of COVID-19 versus 2.5% and 3.0% of delta cases (Table 1).

Table 2 shows VE against delta and omicron infection or hospitalization. Overall, the 1-dose VE was 56.7% (95% CI: 40.7–68.4%) and 20.4% (9.5–30.0%) against delta and omicron infection, respectively.

**Table 2.**
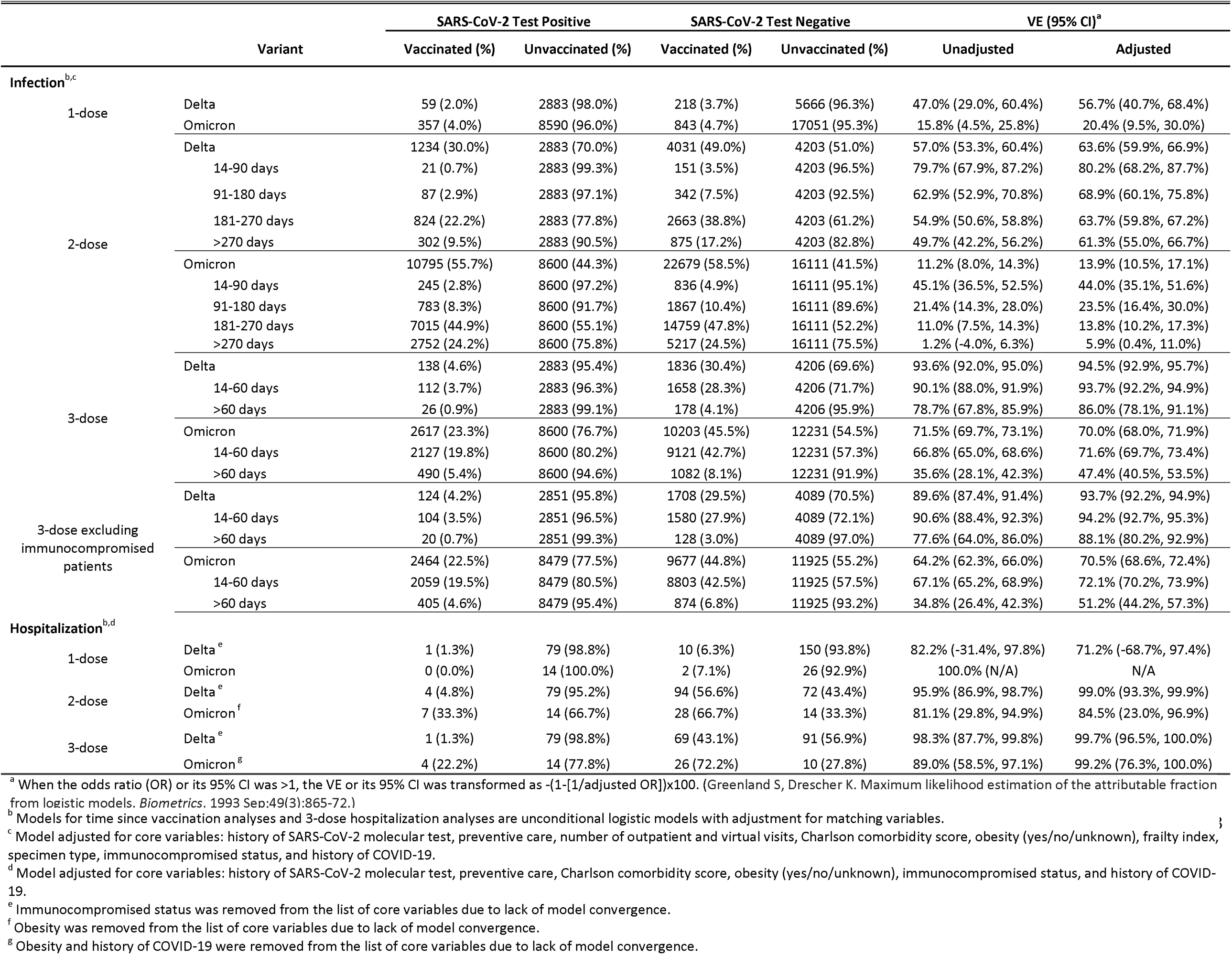
Vaccine effectiveness of mRNA-1273 against infection and hospitalization with delta or omicron variants

In analyses of 2-dose VE against delta infection by time since receipt of dose 2, VE at 14–90 days was 80.2% (68.2–87.7%) and subsequently declined, with VE of 68.9% (60.1–75.8%) at 91–180 days, 63.7% (59.8–67.2%) at 181–270 days and 61.3% (55.0– 66.7%) at >270 days (Table 2, Fig. 2). In comparison, the 2-dose VE against omicron infection was 44.0% (35.1–51.6%) at 14–90 days and declined quickly to 23.5% (16.4– 30.0%) at 91–180 days, 13.8% (10.2–17.3%) at 181–270 days and 5.9% (0.4–11.0%) at >270 days. The 3-dose VE against delta infection was 93.7% (92.2–94.9%) at 14-60 days and 86.0% (78.1–91.1%) at >60 days. However, the 3-dose VE against omicron infection was 71.6% (69.7–73.4%) at 14-60 days and 47.4% (40.5–53.5%) at >60 days. These estimates were similar in analyses that excluded individuals who were immunocompromised, except that the 3-dose VE against omicron infection increased to 51.2% (44.2–57.3%) among immunocompetent individuals at >60 days (Table 2, Fig. 3). The VE of 2 and 3 doses against hospitalization with delta were both ≥99%, while they were 84.5% (23.0–96.9%) and 99.2% (76.3–100.0%) against hospitalization with omicron (Table 2). Notably, all four individuals hospitalized with omicron despite receipt of three mRNA-1273 doses were more than 60 years of age with chronic diseases, and one was also immunocompromised (data not shown).

**Fig. 2:**
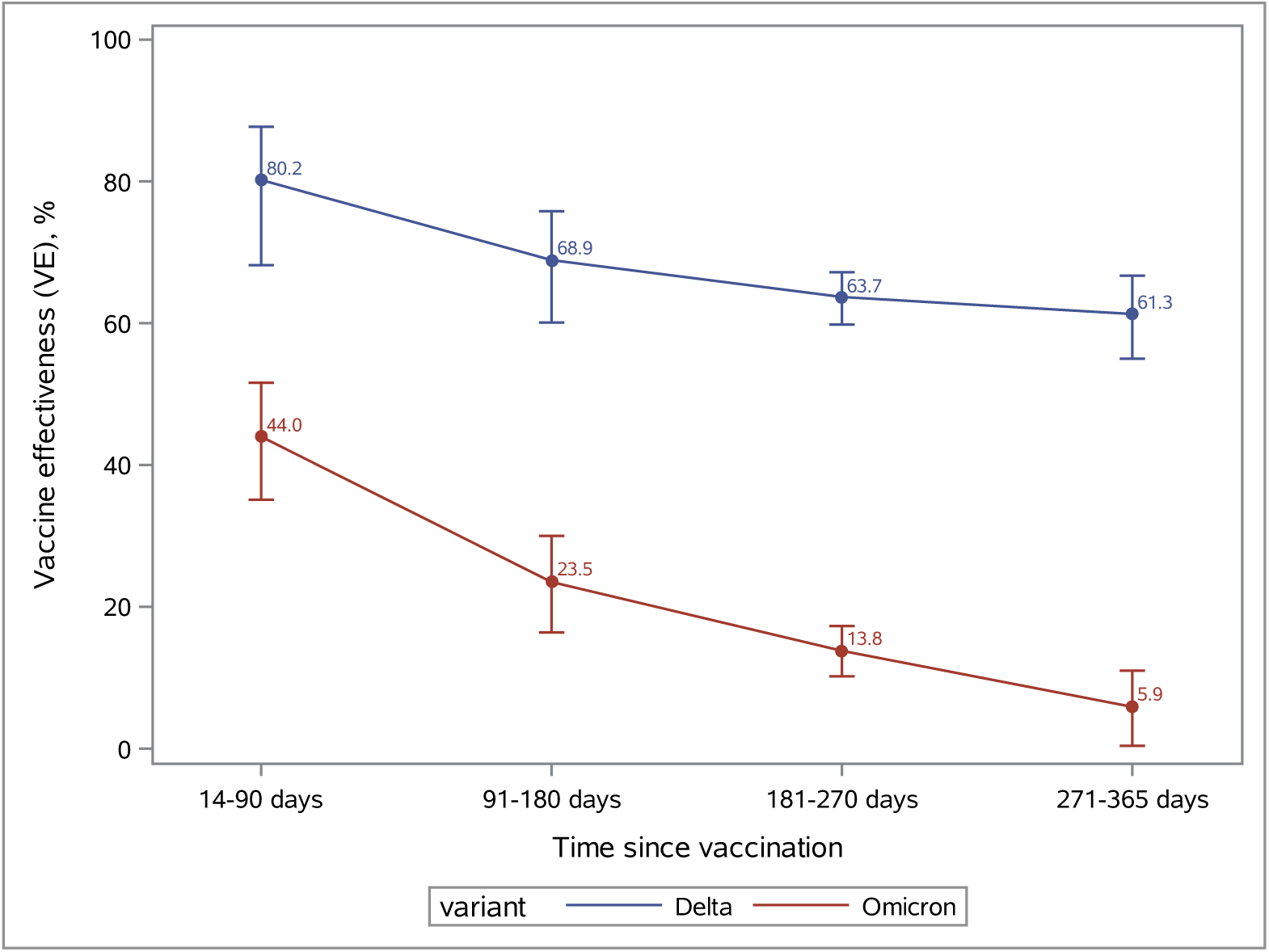
Vaccine effectiveness of 2 doses of mRNA-1273 against omicron and delta variants by time since vaccination. The figure depicts the waning effectiveness of 2 doses of mRNA-1273 vaccine against omicron infection (red line) and delta infection (blue line) within 365 days after receipt of second dose. The vertical bar associated with each point estimate represents the 95% confidence interval of the vaccine effectiveness.

**Fig. 3:**
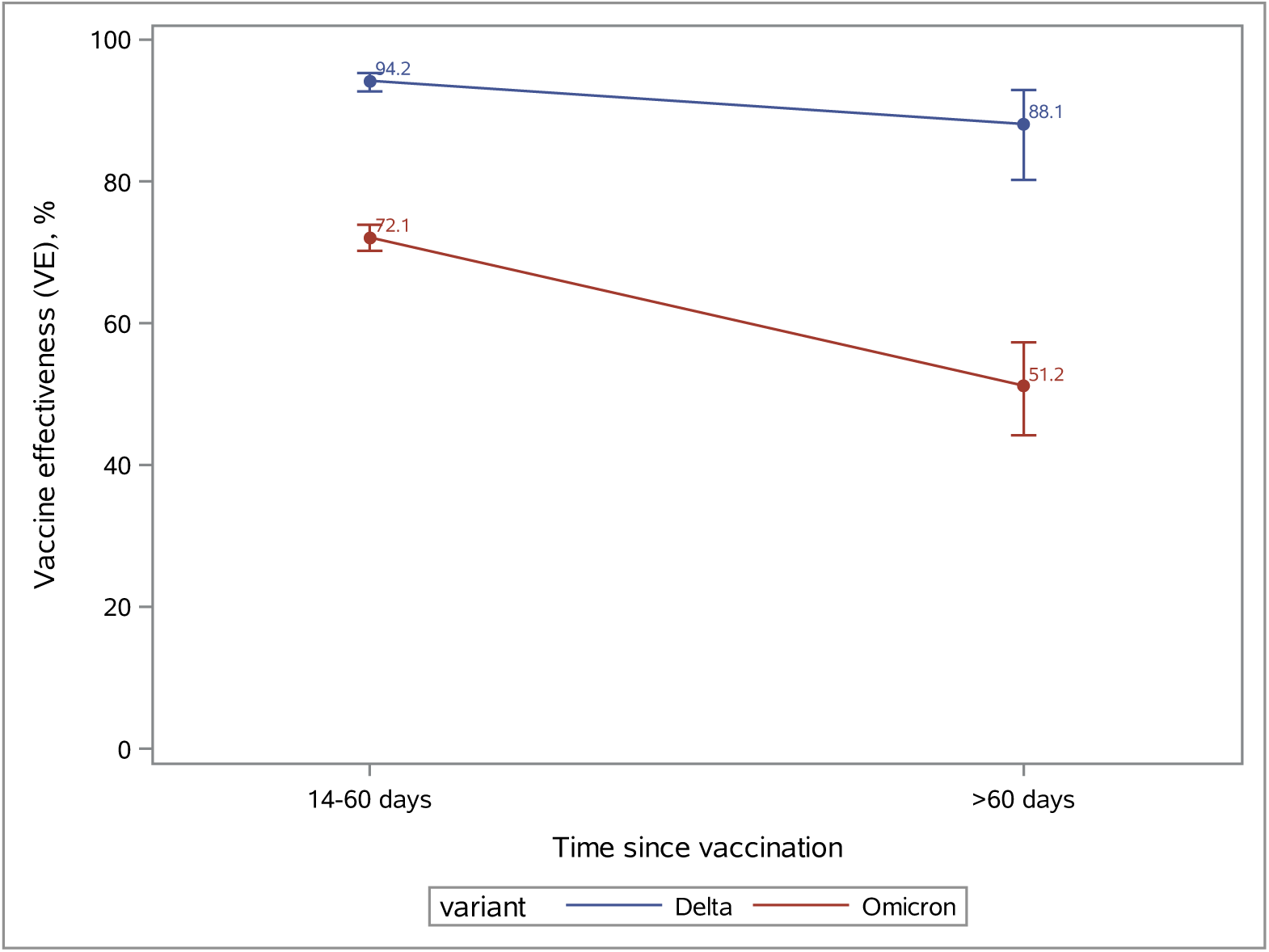
Vaccine effectiveness of 3 doses of mRNA-1273 against omicron and delta variants by time since vaccination among immunocompetent population. The figure depicts the effectiveness of 3 doses of mRNA-1273 vaccine against delta infection (blue line) and omicron infection (red line), comparing effectiveness by time since third dose (14-60 days or >60 days). The vertical bar associated with each point estimate represents the 95% confidence interval of the vaccine effectiveness.

Table 3 presents the 3-dose VE against infection by subgroups. The 3-dose VE against delta infection was >93% across age, sex and race/ethnicity groups but lower in the immunocompromised population (70.6% [31.0–87.5%], p value for interaction <0.001). The 3-dose VE against omicron infection was 70.9% (68.9–72.9%) in those aged <65 years and 64.3% (55.0–71.7%) in those aged ≥65 years, and only 29.4% (0.3–50.0%) in the immunocompromised population compared to 70.5% (68.6–72.4%) in the immunocompetent population (p value for interaction <0.001). The 3-dose VE against omicron infection among those who had no history of COVID-19 was 70.1% (68.0– 72.1%) in those aged <65 years, and 64.5% (54.9–72.1%) in those aged ≥65 years (data not shown).

**Table 3.** Vaccine effectiveness of 3 doses of mRNA-1273 against infection with delta or omicron variants by subgroup

| Variant<sup>a,b</sup> | SARS-CoV-2 Test Positive |  | SARS-CoV-2 Test Negative |  | VE (95% CI) |  | p value for interaction |
| --- | --- | --- | --- | --- | --- | --- | --- |
|  | Vaccinated (%) | Unvaccinated (%) | Vaccinated (%) | Unvaccinated (%) | Unadjusted | Adjusted |  |
| Delta |  |  |  |  |  |  |  |
| Age at specimen collection date |  |  |  |  |  |  | 0.3742 |
| <65 | 94 (3.4%) | 2694 (96.6%) | 1470 (26.4%) | 4106 (73.6%) | 93.3% (91.3%, 94.8%) | 94.3% (92.5%, 95.7%) |  |
| ≥65 | 44 (18.9%) | 189 (81.1%) | 366 (78.5%) | 100 (21.5%) | 95.0% (91.1%, 97.1%) | 96.0% (92.3%, 97.9%) |  |
| Sex |  |  |  |  |  |  | 0.8922 |
| Female | 75 (4.7%) | 1519 (95.3%) | 969 (30.4%) | 2219 (69.6%) | 93.2% (90.7%, 95.0%) | 94.4% (92.2%, 96.0%) |  |
| Male | 63 (4.4%) | 1364 (95.6%) | 867 (30.4%) | 1987 (69.6%) | 94.2% (91.7%, 95.9%) | 94.6% (92.0%, 96.3%) |  |
| Race/ethnicity |  |  |  |  |  |  | 0.1993 |
| Hispanic | 39 (3.0%) | 1240 (97.0%) | 577 (22.6%) | 1981 (77.4%) | 92.4% (88.7%, 94.8%) | 93.1% (89.4%, 95.5%) |  |
| Non-Hispanic and others | 99 (5.7%) | 1643 (94.3%) | 1259 (36.1%) | 2225 (63.9%) | 94.2% (92.2%, 95.7%) | 95.1% (93.2%, 96.4%) |  |
| Immunocompromised status |  |  |  |  |  |  | 0.0002 |
| Yes<sup>c</sup> | 14 (30.4%) | 32 (69.6%) | 128 (52.2%) | 117 (47.8%) | 60.0% (21.4%, 79.7%) | 70.6% (31.0%, 87.5%) |  |
| No | 124 (4.2%) | 2851 (95.8%) | 1708 (29.5%) | 4089 (70.5%) | 89.6% (87.4%, 91.4%) | 93.7% (92.2%, 94.9%) |  |
| Omicron |  |  |  |  |  |  |  |
| Age at specimen collection date |  |  |  |  |  |  | 0.0969 |
| <65 | 1943 (18.9%) | 8335 (81.1%) | 8573 (41.7%) | 11983 (58.3%) | 72.2% (70.4%, 73.9%) | 70.9% (68.9%, 72.9%) |  |
| ≥65 | 674 (71.8%) | 265 (28.2%) | 1630 (86.8%) | 248 (13.2%) | 61.7% (53.2%, 68.6%) | 64.3% (55.0%, 71.7%) |  |
| Sex |  |  |  |  |  |  | 0.9159 |
| Female | 1529 (24.1%) | 4816 (75.9%) | 5862 (46.2%) | 6828 (53.8%) | 70.4% (67.9%, 72.6%) | 70.0% (67.4%, 72.4%) |  |
| Male | 1088 (22.3%) | 3784 (77.7%) | 4341 (44.6%) | 5403 (55.4%) | 72.9% (70.3%, 75.3%) | 70.0% (66.6%, 72.9%) |  |
| Race/ethnicity |  |  |  |  |  |  | 0.0866 |
| Hispanic | 970 (18.9%) | 4157 (81.1%) | 3976 (38.8%) | 6278 (61.2%) | 69.6% (66.7%, 72.2%) | 68.0% (64.6%, 71.0%) |  |
| Non-Hispanic and others | 1647 (27.0%) | 4443 (73.0%) | 6227 (51.1%) | 5953 (48.9%) | 72.8% (70.5%, 74.9%) | 71.4% (68.8%, 73.8%) |  |
| Immunocompromised status |  |  |  |  |  |  | <.0001 |
| Yes | 153 (55.8%) | 121 (44.2%) | 526 (63.2%) | 306 (36.8%) | 26.4% (3.0%, 44.2%) | 29.4% (0.3%, 50.0%) |  |
| No | 2464 (22.5%) | 8479 (77.5%) | 9677 (44.8%) | 11925 (55.2%) | 64.2% (62.3%, 66.0%) | 70.5% (68.6%, 72.4%) |  |

<sup>a</sup> Models for immunocompromised status subgroup analyses are unconditional logistic models with adjustment for matching variables.
<sup>b</sup> Model adjusted for core variables: history of SARS-CoV-2 molecular test, preventive care, number of outpatient and virtual visits, Charlson comorbidity score, obesity (yes/no/unknown), frailty index, specimen type, immunocompromised status, and history of COVID-19.
<sup>c</sup> Number of outpatient and virtual visits was removed from the list of core variables due to lack of model convergence.

## Discussion

We evaluated the effectiveness of mRNA-1273 against the highly mutated omicron variant in a socio-demographically diverse population in a real-world setting. Between December 6, 2021, and December 31, 2021, the rapidly increasing proportion of omicron-positive specimens indicated unprecedented transmissibility and raised concerns over protection conferred by currently authorized or licensed COVID-19 vaccines. Our study demonstrates that while VE of 2 doses of mRNA-1273 against delta infection is high and wanes slowly, consistent with our previous findings,^6,14^ the 2-dose VE against omicron infection is inadequate, providing only modest protection of 44.0% within 3 months of vaccination and diminishing quickly thereafter. In addition, while the 3-dose VE against delta infection is high and durable, that against omicron is lower. Nevertheless, the average point estimate (>50%) and lower bound of the 95% CI (>30%) still meet the US FDA criteria for emergency use authorization for COVID-19 vaccines.^15^ Also, this VE is similar to the 2-dose vaccine efficacy against asymptomatic infection observed in the phase 3 clinical trial (63.0% [56.6–68.5%]).^16^ The VE of 3 doses of mRNA-1273 against omicron infection is poor among individuals who are immunocompromised. While 2-dose VE against hospitalization with omicron is lower compared to that with delta, 3-dose VE is nearly 100% against hospitalization with either variant. Although additional study is needed, these findings suggest that third (booster) doses may be needed <6 months after dose 2 in immunocompetent individuals and that 3 doses may be inadequate to protect against omicron infection in individuals who are immunocompromised. Furthermore, the data indicate a potential need for periodic adjustment of vaccines to target circulating variants that have evolved to escape current vaccine-induced immunity.

While there are limited prior data on VE of 2 or 3 doses of mRNA-1273 vaccine against infection or hospitalization with omicron, a preliminary analysis from Denmark found an initial VE of 2 doses of mRNA-1273 against omicron infection of 36.7% that waned quickly, similar to our findings.^17^ An early report by Andrews et al^18^ found waning of 2-dose protection with an initial VE of 2 doses of BNT162b2 against symptomatic omicron infection of 88% (65.9–95.8%) 2–9 weeks after dose 2 that declined to 34–37% (95% CIs ranging from –5 to 59.6%) 15 or more weeks after dose 2, but increased to 75.5% (56.1–86.3%) a median of 41 days (range 14–72 days) after a BNT162b2 booster. Collie et al^19^ found that the VE of 2 doses of BNT162b2 against hospitalization during a proxy omicron period was 70% at least 14 days after receipt of dose 2. In England, after a primary course of BNT162b2 vaccine, VE against omicron infection was initially 70% after a BNT162b2 booster, dropping to 45% after ≥10 weeks, but stayed around 70– 75% for up to 9 weeks after an mRNA-1273 booster.^12^

A growing number of reports indicate that omicron disease is less severe than delta disease, resulting in a lower risk of hospitalization.^1,20^ This may reflect greater replication of omicron in the upper versus lower respiratory tract, which may also contribute to more efficient transmission, resulting in increased absolute^21^ numbers of hospitalizations. Booster vaccination has the potential to decrease hospital burden and improve outcomes.^22^ While the sample size and follow-up period were not sufficient in our study or other studies to assess potential waning VE against hospitalization with omicron, our results of waning VE against omicron infection after dose 3 of mRNA-1273 underscores the importance of monitoring VE against hospitalization with omicron. This study provides novel data complementing recent reports of the effectiveness of other COVID-19 vaccines against omicron infection and has several strengths and limitations.^14,23^ First, the results of our test-negative case-control study may not be generalizable to people who are not tested, including those with milder symptoms who may not pursue testing. While there is a variety of reasons for testing that may introduce biases, we attempted to reduce these biases by accounting for sociodemographic characteristics, prior health care utilization, SARS-CoV-2 testing and comorbidities in the models. Although potential residual confounding or detection bias could remain, they were not likely to reverse the conclusion of the study. While misclassification of disease status was a potential source of bias, we used a highly specific and sensitive RT-PCR test that likely minimized misclassification and also enabled us to monitor variant proportions through whole genome sequencing and SGTF analysis. Similarly, misclassification of vaccination status was possible but likely minimal and non-differential with respect to disease status. KPSC electronic vaccination records that captured all vaccine administrations given at KPSC were also updated daily with vaccine administration data from the California Immunization Registry to which all facilities were required by law to report COVID-19 vaccine administrations within 24 hours of administration. Second, we considered all SGTF specimens as omicron, as our validation samples using whole genome sequencing showed high agreement. Our rate of SGTF closely mirrored regional trends in omicron emergence from the Centers for Disease Control and Prevention (CDC).^13^ Notably, prior to the study, delta accounted for 99% of variants for a few months prior to the emergence of omicron in Southern California. Furthermore, during the study interval, delta and omicron accounted for >99% of variants, and the BA.2 sub-lineage of omicron was not detected among any of the 1,383 specimens sequenced in this study. Therefore, virtually all variants exhibiting SGTF were omicron while those without SGTF were delta during the study interval. Third, this study was representative of a large, diverse racial, ethnic, and socioeconomic population in Southern California but may be less representative of other populations. However, analysis of the effectiveness of mRNA-1273 against delta and omicron in parallel provided an internal comparator that put results in context.^14^ Fourth, some individuals who were immunocompetent and who received a third dose before the October 21, 2021, Advisory Committee on Immunization Practices (ACIP) recommendation may have received a 100-µg dose rather than a 50-µg booster dose of mRNA-1273. However, we were not able to clearly assess the difference, as dosage information was not available from external vaccination records. Fifth, the number of hospitalized individuals included was too small to draw definitive conclusions regarding VE and durability of 3 doses in preventing hospitalization. Long-term follow-up is needed to evaluate the durability of both 100-µg and 50-µg booster doses in preventing infection and hospitalization. Sixth, we did not evaluate VE against symptomatic or asymptomatic infection. However, we did find higher VE against COVID-19 hospitalization. Aside from the saliva tests that were only collected in asymptomatic individuals, information on whether infections were symptomatic or asymptomatic was not readily available in structured data. For future analyses, we plan to apply a natural language processing algorithm to clinical notes to differentiate symptomatic from asymptomatic SARS-CoV-2 infections. Finally, caution should be taken when interpreting waning VE over time as some confidence intervals overlapped, and heterogenous composition of the vaccinated population over time could potentially contribute to varying estimates as well. Among the populations first prioritized for vaccination, the most clinically vulnerable individuals may have contributed to over-estimates in waning, although this may have potentially been offset to some extent by health care workers with less waning. Furthermore, early vaccine adopters may have implemented risk-avoidance behaviors that put them at a lower risk of infection.

In conclusion, this study of mRNA-1273 found waning 2-dose but high 3-dose VE against delta infection and lower 2-dose and 3-dose VE against omicron infection. The 2-dose VE against hospitalization with omicron is lower than with delta but the 3-dose VE against hospitalization with either variant is excellent. Protection against omicron infection wanes within 3 months after dose 2, suggesting that a shorter interval between second and booster doses may be beneficial. Lack of protection against omicron infection in the immunocompromised population underscores the importance of monitoring the effectiveness of the recommended fourth dose (booster) for this population. Continued monitoring of VE against omicron infection and hospitalization in immunocompetent and immunocompromised individuals and surveillance for the emergence of newer SARS-CoV-2 variants are warranted to inform future vaccination strategies.

## Data Availability

Individual-level data reported in this study are not publicly shared. Upon request, and subject to review, KPSC may provide the deidentified aggregate-level data that support the findings of this study. Deidentified data (including participant data as applicable) may be shared upon approval of an analysis proposal and a signed data access agreement.

## Acknowledgments

This study was funded by Moderna, Inc. Manuscript authors, including those employed by the funder, contributed to the conceptualization, design, interpretation of data, decision to publish, and preparation of the manuscript. Medical writing and editorial assistance were provided by Srividya Ramachandran, PhD, and Jared Mackenzie, PhD, of MEDiSTRAVA in accordance with Good Publication Practice (GPP3) guidelines, funded by Moderna, Inc., and under the direction of the authors. Laboratory and database support were provided at KPSC by Lee Childs, Julie Stern, Joy Gelfond, Radha Bathala, Kourtney Kottmann, Ana Acevedo, Elmer Ayala, Samantha Quinones, Samantha Baluyot, Errol Lopez and Don McCarthy. The authors thank the KPSC Lab Leadership and Technician Team for their support of this study. The authors would like to acknowledge Helix OpCo, LLC, for their whole genome sequencing of SARS-CoV-2 specimens. The authors thank the patients of Kaiser Permanente for their partnership with us to improve their health. Their information, collected through our electronic health record systems, leads to findings that help us improve care for our members and can be shared with the larger community. Julie Vanas, Moderna, Inc., provided critical operations support and Yamuna Paila, Moderna, Inc., provided critical input on specimen sequencing.

## Author contributions

H.F.T, B.K.A., L.S.S, L.Q, K.J.B, and C.A.T were involved in the study concept and design, as well as acquisition, analysis, or interpretation of data. H.F.T and B.K.A drafted the manuscript. Y.L, L.S.S, C.A.T, Y.T, K.J.B, J.E.T, A.F, J.H.K, G.S.L, S.K.C, H.S.T, M.A, and L.Q critically revised the manuscript for important intellectual content. L.Q, Y.L, Y.T, and J.E.T conducted the statistical analyses. L.S.S, C.A.T, G.S.L, M.A, S.K.C, and H.S.T provided administrative, technical, or material support. C.A.T and H.F.T obtained funding and provided supervision.

## Online Methods

### Study setting

KPSC is an integrated health care system that provides care to more than 4.6 million socio-demographically diverse health plan members at 15 hospitals and associated medical offices across Southern California. Comprehensive EHRs used for this study included information on demographics, immunizations, diagnoses, laboratory tests, procedures and pharmacy records. KPSC began administering mRNA-1273 on 12/18/2020. Outside COVID-19 vaccinations were imported into members’ EHRs daily from external sources, including the California Immunization Registry, Care Everywhere (system on the Epic EHR platform that allows health care systems to exchange members’ medical information), claims (eg, retail pharmacies) and self-report by members (with valid documentation).

### Laboratory methods

Molecular diagnostic testing for SARS-CoV-2 is available to members who request it for any reason, before procedures and hospital admissions, with and without symptoms. Specimens were primarily collected using nasopharyngeal/oropharyngeal swabs (for symptomatic or asymptomatic individuals) or saliva (for asymptomatic individuals). Specimens were tested using RT-PCR TaqPath COVID-19 High-Throughput Combo Kit (Thermo Fisher Scientific). SGTF was defined as a RT-PCR test in which N and ORF1ab genes were detected (Ct values <37), but S gene was not detected. Specimens with SGTF were considered to be omicron, whereas positive specimens without SGTF were considered to be delta.

A random sample of SARS-CoV-2 positive specimens were sent for whole genome sequencing (WGS). Details have been described in our previous publication.^14^ The SGTF data were compared against WGS results to assess their validity in differentiating variants.

### Study design

A test-negative case-control study design was used in which individuals testing positive for SARS-CoV-2 were defined as cases and individuals testing negative were defined as controls; this design is purported to reduce bias associated with confounding by health care-seeking behavior and misclassification of cases.^24^ In this study, cases included individuals who tested positive by the RT-PCR TaqPath COVID-19 kit, had specimens collected between 12/6/2021 and 12/31/2021, were aged ≥18 years, and had ≥12 months of KPSC membership before the specimen collection date (for accurate ascertainment of exposure status and covariates). Individuals were excluded if they received a COVID-19 vaccine other than mRNA-1273, any dose of mRNA-1273 <14 days before the specimen collection date, 2 or 3 doses of mRNA-1273<24 days apart from previous dose or >3 doses of mRNA-1273 prior to the specimen collection date. Additional exclusions included a positive SARS-CoV-2 test or COVID-19 diagnosis code ≤90 days before the specimen collection date. COVID-19 hospitalization included hospitalization with a SARS-CoV-2–positive test or hospitalization ≤7 days after a SARS-CoV-2–positive test. COVID-19 hospitalization was confirmed by manual chart review conducted by a physician investigator (B.K.A.) to verify the presence of severe COVID-19 symptoms.

Controls included all individuals who tested negative with specimens collected between 12/6/2021 and 12/31/2021, were aged ≥18 years, and had ≥12 months of KPSC membership before the specimen collection date. Randomly sampled controls were 2:1 matched to cases by age (18–44 years, 45–64 years, 65–74 years and ≥75 years), sex, race/ethnicity (non-Hispanic White, non-Hispanic Black, Hispanic, non-Hispanic Asian and other/unknown) and specimen collection date. Matching was conducted separately for the 1-, 2-, and 3-dose VE analysis. To accommodate variation in real-world practice, analyses did not require dose 3 to be ≥6 months from dose 2, as some members received dose 3 at a shorter interval in this study.

### Exposure

The exposure of interest was 1, 2 or 3 doses of mRNA-1273. Dose 3 in this analysis included both the 100-µg additional primary dose in individuals who were immunocompromised, as well as the 50-µg and 100-ug booster dose in adults.

### Covariates

A comprehensive list of pre-specified potential confounders were identified a priori based on the literature. Demographic and clinical covariates were extracted from EHRs.^14^ Variables assessed included socioeconomic status (Medicaid, neighborhood median household income), medical center area, pregnancy status, KPSC physician/employee status, smoking, body mass index (BMI), Charlson comorbidity score, autoimmune conditions, chronic diseases (kidney, heart, lung, and liver disease and diabetes), frailty index and immunocompromised status (HIV/AIDS, leukemia/lymphoma, congenital and other immunodeficiencies, asplenia/hyposplenia, hematopoietic stem cell and organ transplant, and/or immunosuppressant medications). To account for potential differences in care-seeking or test-seeking behaviors, the following variables were also assessed: health care utilization (virtual, outpatient, emergency department and inpatient encounters), preventive care (other vaccinations, screenings and wellness visits), history of SARS-CoV-2 molecular test performed from 3/1/2020 to specimen collection date (irrespective of result) and history of COVID-19 (positive SARS-CoV-2 molecular test or a COVID-19 diagnosis code) from 3/1/2020 to specimen collection date.

### Statistical analyses

Characteristics of cases and controls for each analysis were compared by using the χ^2^ test or Fisher exact test for categorical variables and two-sample *t* test or Wilcoxon rank sum test for continuous variables. Absolute standardized difference was calculated to assess the balance of covariates. The distribution of variant type by vaccination status was tabulated. Conditional logistic regression was used to estimate the adjusted odds ratios (OR) and 95% confidence intervals (CI) for vaccination against infection and hospitalization with delta or omicron. In order to harmonize the covariates adjusted across different models so that estimates were comparable, we selected two sets of core variables to be included in all models, one set for infection models and one set for hospitalization models. The selection of core variables was based on prior knowledge, potential associations with infection/hospitalization, and model parsimony, allowing us to control for test/care seeking behavior, general health status, test type, and immunity. For the infection models, the core variables included history of SARS-CoV-2 molecular test, preventive care, number of outpatient and virtual visits, Charlson comorbidity score, obesity (BMI≥30), frailty index, specimen type, immunocompromised status, and history of COVID-19. For the hospitalization models, the core variables included history of SARS-CoV-2 molecular test, preventive care, Charlson comorbidity score, obesity (BMI≥30), immunocompromised status, and history of COVID-19. Unconditional logistic regression with additional adjustment of matching factors in the model was used when matched sets needed to be broken for certain subgroup analyses or when the conditional model failed to converge. VE (%) was calculated as (1–adjusted OR)×100.

We also assessed 2-dose and 3-dose VE against delta or omicron infection by time since receipt of mRNA-1273 dose 2 or 3 (for 2-dose VE: 14-90 days, 91-180 days, 181-270 days, and >270 days; for 3-dose VE: 14-60 days and >60 days). As more immunocompromised persons might have received dose 3 before the October 21, 2021 ACIP booster dose recommendation ^25,26^, we conducted a separate analysis that excluded individuals who were immunocompromised to assess durability of protection of 3 doses in immunocompetent individuals. We also evaluated 3-dose VE in select subgroups, including by age (<65, ≥65 years), sex, race/ethnicity (Hispanic, Non-Hispanic and others) and immunocompromised status (yes, no). The difference between subgroups was tested by including an interaction term for subgroup and vaccination in the model. As VE in individuals with a history of COVID-19 is different from those without,^6^ we also evaluated 3-dose VE against omicron infection, stratified by age (<65 years and ≥65 years), among individuals with no history of COVID-19. SAS 9.4 was used for analyses.

The study was approved by KPSC Institutional Review Board. All study staff with access to protected health information were trained in procedures to protect the confidentiality of KPSC member data. A waiver of informed consent was obtained as this is an observational study of authorized and recommended Moderna COVID-19 vaccine administered in the course of routine clinical care. To facilitate the conduct of this study, a waiver was obtained for written HIPAA authorization for research involving use of the EHR.

## Data availability

Individual-level data reported in this study are not publicly shared. Upon request, and subject to review, KPSC may provide the deidentified aggregate-level data that support the findings of this study. Deidentified data may be shared upon approval of an analysis proposal and a signed data access agreement.

## Code availability

Standard epidemiological analyses were conducted using standard commands in SAS 9.4 (SAS Institute, Cary NC). The commands/code are in the process of being deposited into GitHub.

