## Supplementary Material for "Effectiveness of mRNA-1273 against SARS-CoV-2 omicron and delta variants"

Supplementary Tables

**Authors:** Hung Fu Tseng, PhD,[1,2,*] Bradley K. Ackerson, MD,[1] Yi Luo, PhD,[1] Lina S. Sy, MPH,[1] Carla A. Talarico, PhD,[3] Yun Tian, MS,[1] Katia J. Bruxvoort, PhD,[4] Julia E. Tubert, MPH,[1] Ana Florea, PhD,[1] Jennifer H. Ku, PhD,[1] Gina S. Lee, MPH,[1] Soon Kyu Choi, MPH,[1] Harpreet S. Takhar, MPH,[1] Michael Aragones, MD,[1] Lei Qian, PhD[1]

**Affiliations:** [1]Department of Research and Evaluation, Kaiser Permanente Southern California, 100 S. Los Robles Ave., Pasadena, CA, 91101, USA; [2]Department of Health Systems Science, Kaiser Permanente Bernard J. Tyson School of Medicine, 98 S. Los Robles Ave., Pasadena, CA, 91101, USA; [3]Moderna Inc., 200 Technology Square, Cambridge, MA, 02139, USA; [4]Department of Epidemiology, University of Alabama at Birmingham, 1665 University Blvd., Birmingham, AL, 35233, USA

Supplementary Table 1. Characteristics of SARS-CoV-2 cases and controls, by variant (1-dose analysis)

| | Delta | | | Omicron | | |
|---|---|---|---|---|---|---|
| | Test Positive Cases N=2942 | Test Negative Controls N=5884 | p-value/ASD | Test Positive Cases N=8947 | Test Negative Controls N=17894 | p-value/ASD |
| Age at specimen collection date, years | | | 0.91 / 0.01 | | | <0.01 / 0.04 |
| mean (sd) | 41.24 (14.29) | 41.33 (14.36) | | 37.40 (13.06) | 37.92 (13.25) | |
| median | 39 | 39 | | 35 | 36 | |
| Q1, Q3 | 31, 51 | 31, 51 | | 27, 46 | 28, 46 | |
| min, max | 18, 90 | 18, 101 | | 18, 93 | 18, 96 | |
| Age at specimen collection date, years, n (%) | | | N/A | | | N/A |
| 18-44 | 1844 (62.7%) | 3688 (62.7%) | | 6477 (72.4%) | 12954 (72.4%) | |
| 45-64 | 904 (30.7%) | 1808 (30.7%) | | 2188 (24.5%) | 4376 (24.5%) | |
| 65-74 | 153 (5.2%) | 306 (5.2%) | | 238 (2.7%) | 476 (2.7%) | |
| ≥75 | 41 (1.4%) | 82 (1.4%) | | 44 (0.5%) | 88 (0.5%) | |
| Sex, n (%) | | | N/A | | | N/A |
| Female | 1550 (52.7%) | 3100 (52.7%) | | 4996 (55.8%) | 9992 (55.8%) | |
| Male | 1392 (47.3%) | 2784 (47.3%) | | 3951 (44.2%) | 7902 (44.2%) | |
| Race/Ethnicity, n (%) | | | N/A | | | N/A |
| Non-Hispanic White | 1159 (39.4%) | 2318 (39.4%) | | 2526 (28.2%) | 5052 (28.2%) | |
| Non-Hispanic Black | 181 (6.2%) | 362 (6.2%) | | 961 (10.7%) | 1922 (10.7%) | |
| Hispanic | 1265 (43.0%) | 2530 (43.0%) | | 4319 (48.3%) | 8638 (48.3%) | |
| Non-Hispanic Asian | 97 (3.3%) | 194 (3.3%) | | 387 (4.3%) | 774 (4.3%) | |
| Other/Unknown | 240 (8.2%) | 480 (8.2%) | | 754 (8.4%) | 1508 (8.4%) | |
| Body mass index[b], n (%) | | | <0.01 / 0.15 | | | <0.01 / 0.08 |
| <18.5 | 20 (0.7%) | 71 (1.2%) | | 105 (1.2%) | 224 (1.3%) | |
| 18.5 - <25 | 545 (18.5%) | 1273 (21.6%) | | 1730 (19.3%) | 3645 (20.4%) | |
| 25 - <30 | 740 (25.2%) | 1591 (27.0%) | | 2377 (26.6%) | 4609 (25.8%) | |
| 30 - <35 | 590 (20.1%) | 1084 (18.4%) | | 1655 (18.5%) | 3503 (19.6%) | |
| 35 - <40 | 295 (10.0%) | 607 (10.3%) | | 837 (9.4%) | 1806 (10.1%) | |
| 40 - <45 | 117 (4.0%) | 247 (4.2%) | | 393 (4.4%) | 795 (4.4%) | |
| ≥45 | 66 (2.2%) | 147 (2.5%) | | 245 (2.7%) | 539 (3.0%) | |
| Unknown | 569 (19.3%) | 864 (14.7%) | | 1605 (17.9%) | 2773 (15.5%) | |
| Smoking[b], n (%) | | | <0.01 / 0.12 | | | <0.01 / 0.07 |
| No | 1981 (67.3%) | 4103 (69.7%) | | 6375 (71.3%) | 12709 (71.0%) | |
| Yes | 484 (16.5%) | 1068 (18.2%) | | 1268 (14.2%) | 2883 (16.1%) | |
| Unknown | 477 (16.2%) | 713 (12.1%) | | 1304 (14.6%) | 2302 (12.9%) | |
| Charlson comorbidity score[a], n (%) | | | 0.01 / 0.07 | | | <0.01 / 0.13 |
| 0 | 2443 (83.0%) | 4760 (80.9%) | | 7721 (86.3%) | 14725 (82.3%) | |
| 1 | 316 (10.7%) | 652 (11.1%) | | 866 (9.7%) | 2001 (11.2%) | |
| ≥2 | 183 (6.2%) | 472 (8.0%) | | 360 (4.0%) | 1168 (6.5%) | |

| | | | | | | |
|---|---|---|---|---|---|---|
| Frailty index[a], n (%) | | | <0.01 / 0.17 | | | <0.01 / 0.13 |
| Quartile 1 | 731 (24.8%) | 1474 (25.1%) | | 2163 (24.2%) | 4258 (23.8%) | |
| Quartile 2 | 856 (29.1%) | 1350 (22.9%) | | 2571 (28.7%) | 4424 (24.7%) | |
| Quartile 3 | 728 (24.7%) | 1481 (25.2%) | | 2272 (25.4%) | 4442 (24.8%) | |
| Quartile 4 (most frail) | 627 (21.3%) | 1579 (26.8%) | | 1941 (21.7%) | 4770 (26.7%) | |
| Chronic diseases[a], n (%) | | | | | | |
| Kidney disease | 41 (1.4%) | 117 (2.0%) | 0.05 / 0.05 | 62 (0.7%) | 289 (1.6%) | <0.01 / 0.09 |
| Heart disease | 34 (1.2%) | 108 (1.8%) | 0.02 / 0.06 | 59 (0.7%) | 232 (1.3%) | <0.01 / 0.06 |
| Lung disease | 193 (6.6%) | 461 (7.8%) | 0.03 / 0.05 | 531 (5.9%) | 1393 (7.8%) | <0.01 / 0.07 |
| Liver disease | 68 (2.3%) | 162 (2.8%) | 0.22 / 0.03 | 175 (2.0%) | 483 (2.7%) | <0.01 / 0.05 |
| Diabetes | 170 (5.8%) | 386 (6.6%) | 0.15 / 0.03 | 398 (4.4%) | 1064 (5.9%) | <0.01 / 0.07 |
| Immunocompromised, n (%) | 32 (1.1%) | 173 (2.9%) | <0.01 / 0.13 | 131 (1.5%) | 488 (2.7%) | <0.01 / 0.09 |
| HIV/AIDS | 2 | 4 | | 12 | 30 | |
| Leukemia/lymphoma, congenital and other immunodeficiencies, asplenia/hyposplenia | 12 | 62 | | 44 | 150 | |
| Hematopoietic stem cell transplantation/ organ transplant | 2 | 6 | | 7 | 29 | |
| Immunosuppressant medications | 18 | 120 | | 83 | 342 | |
| Autoimmune conditions[a], n (%) | 55 (1.9%) | 160 (2.7%) | 0.01 / 0.06 | 146 (1.6%) | 373 (2.1%) | 0.01 / 0.03 |
| Rheumatoid arthritis | 13 | 54 | | 49 | 165 | |
| Inflammatory bowel disease | 15 | 50 | | 42 | 92 | |
| Psoriasis and psoriatic arthritis | 23 | 53 | | 44 | 94 | |
| Multiple sclerosis | 4 | 8 | | 12 | 24 | |
| Systemic lupus erythematosus | 3 | 12 | | 19 | 55 | |
| Pregnant at specimen collection date, n (%) | 57 (1.9%) | 203 (3.5%) | <0.01 / 0.09 | 195 (2.2%) | 774 (4.3%) | <0.01 / 0.12 |
| 1st trimester | 16 | 28 | | 33 | 80 | |
| 2nd trimester | 19 | 48 | | 69 | 174 | |
| 3rd trimester | 22 | 127 | | 93 | 520 | |
| History of COVID-19[c], n (%) | 79 (2.7%) | 1354 (23.0%) | <0.01 / 0.64 | 1622 (18.1%) | 4473 (25.0%) | <0.01 / 0.17 |
| History of SARS-CoV-2 molecular test[c], n (%) | 1886 (64.1%) | 4684 (79.6%) | <0.01 / 0.35 | 6563 (73.4%) | 13347 (74.6%) | 0.03 / 0.03 |
| Number of outpatient and virtual visits[a], n (%) | | | <0.01 / 0.29 | | | <0.01 / 0.19 |
| 0 | 460 (15.6%) | 586 (10.0%) | | 1196 (13.4%) | 1920 (10.7%) | |
| 1-4 | 1123 (38.2%) | 1823 (31.0%) | | 3460 (38.7%) | 5990 (33.5%) | |
| 5-10 | 711 (24.2%) | 1593 (27.1%) | | 2275 (25.4%) | 4596 (25.7%) | |
| ≥11 | 648 (22.0%) | 1882 (32.0%) | | 2016 (22.5%) | 5388 (30.1%) | |
| Number of Emergency Department visits[a], n (%) | | | <0.01 / 0.18 | | | <0.01 / 0.14 |
| 0 | 2513 (85.4%) | 4642 (78.9%) | | 7449 (83.3%) | 14106 (78.8%) | |
| 1 | 308 (10.5%) | 830 (14.1%) | | 1111 (12.4%) | 2506 (14.0%) | |

| | | | | | | |
|---|---|---|---|---|---|---|
| ≥2 | 121 (4.1%) | 412 (7.0%) | | 387 (4.3%) | 1282 (7.2%) | |
| Number of hospitalizations[a], n (%) | | | 0.02 / 0.07 | | | <0.01 / 0.11 |
| 0 | 2797 (95.1%) | 5512 (93.7%) | | 8561 (95.7%) | 16710 (93.4%) | |
| 1 | 118 (4.0%) | 285 (4.8%) | | 340 (3.8%) | 948 (5.3%) | |
| ≥2 | 27 (0.9%) | 87 (1.5%) | | 46 (0.5%) | 236 (1.3%) | |
| Preventive care[a], n(%) | 1355 (46.1%) | 2965 (50.4%) | <0.01 / 0.09 | 3971 (44.4%) | 8578 (47.9%) | <0.01 / 0.07 |
| Medicaid, n (%) | 303 (10.3%) | 684 (11.6%) | 0.06 / 0.04 | 1077 (12.0%) | 2495 (13.9%) | <0.01 / 0.06 |
| Neighborhood median household income, n(%) | | | 0.04 / 0.07 | | | <0.01 / 0.06 |
| < $40,000 | 127 (4.3%) | 273 (4.6%) | | 387 (4.3%) | 948 (5.3%) | |
| $40,000-$59,999 | 490 (16.7%) | 1132 (19.2%) | | 1789 (20.0%) | 3820 (21.3%) | |
| $60,000-$79,999 | 799 (27.2%) | 1515 (25.7%) | | 2447 (27.3%) | 4724 (26.4%) | |
| $80,000+ | 1524 (51.8%) | 2959 (50.3%) | | 4307 (48.1%) | 8373 (46.8%) | |
| Unknown | 2 (0.1%) | 5 (0.1%) | | 17 (0.2%) | 29 (0.2%) | |
| KPSC physician/employee, n (%) | 75 (2.5%) | 533 (9.1%) | <0.01 / 0.28 | 275 (3.1%) | 466 (2.6%) | 0.03 / 0.03 |
| Specimen type, n (%) | | | <0.01 / 0.55 | | | <0.01 / 0.21 |
| Nasopharyngeal/oropharyngeal swab | 2541 (86.4%) | 3724 (63.3%) | | 7599 (84.9%) | 13730 (76.7%) | |
| Saliva | 401 (13.6%) | 2160 (36.7%) | | 1348 (15.1%) | 4164 (23.3%) | |

[a] Defined in the one year prior to specimen collection date

[b] Defined in the two years prior to specimen collection date

[c] Defined based on all available medical records from March 1, 2020 to specimen collection date

Medical center area not shown. There were differences in the distribution of the vaccinated and unvaccinated individuals across the 19 medical center areas.

N/A = not applicable

### Supplementary Table 2. Full model for 1-dose VE against delta variant infection

**Odds Ratio Estimates**

| Effect | Point Estimate | 95% Wald Confidence Interval | |
|---|---|---|---|
| Vaccination status, Vaccinated vs Unvaccinated | 0.433 | 0.316 | 0.593 |
| History of SARS-CoV-2 molecular test, No vs Yes | 1.347 | 1.201 | 1.512 |
| Preventive Care, No vs Yes | 1.12 | 0.995 | 1.261 |
| Number of outpatient and virtual visits, 0 vs >=11 | 1.581 | 1.236 | 2.022 |
| Number of outpatient and virtual visits, 1-4 vs >=11 | 1.443 | 1.236 | 1.686 |
| Number of outpatient and virtual visits, 5-10 vs >=11 | 1.151 | 0.991 | 1.336 |
| Obese, No vs Yes | 0.857 | 0.764 | 0.961 |
| Obese, Unknown vs Yes | 0.914 | 0.768 | 1.088 |
| Charlson comorbidity score, 0 vs >=2 | 0.969 | 0.766 | 1.226 |
| Charlson comorbidity score, 1 vs >=2 | 1.117 | 0.866 | 1.443 |
| Frailty, Quartile 1 vs Quartile 4 | 1.227 | 1.04 | 1.448 |
| Frailty, Quartile 2 vs Quartile 4 | 1.212 | 0.999 | 1.469 |
| Frailty, Quartile 3 vs Quartile 4 | 1.2 | 1.023 | 1.407 |
| Specimen type, Nasopharyngeal/oropharyngeal swab vs Saliva | 3.657 | 3.206 | 4.172 |
| Immunocompromised status, No vs Yes | 2.479 | 1.622 | 3.789 |
| History of COVID-19 diagnosis, No vs Yes | 10.064 | 7.838 | 12.922 |

### Supplementary Table 3. Full model for 1-dose VE against omicron variant infection

| Odds Ratio Estimates | | | |
|---|---|---|---|
| Effect | Point Estimate | 95% Wald Confidence Interval | |
| Vaccination status, Vaccinated vs Unvaccinated | 0.796 | 0.7 | 0.905 |
| History of SARS-CoV-2 molecular test, No vs Yes | 0.849 | 0.797 | 0.904 |
| Preventive Care, No vs Yes | 1.067 | 1.004 | 1.134 |
| Number of outpatient and virtual visits, 0 vs >=11 | 1.546 | 1.362 | 1.754 |
| Number of outpatient and virtual visits, 1-4 vs >=11 | 1.438 | 1.328 | 1.558 |
| Number of outpatient and virtual visits, 5-10 vs >=11 | 1.265 | 1.172 | 1.366 |
| Obese, No vs Yes | 1.014 | 0.956 | 1.075 |
| Obese, Unknown vs Yes | 0.984 | 0.9 | 1.076 |
| Charlson comorbidity score, 0 vs >=2 | 1.394 | 1.212 | 1.604 |
| Charlson comorbidity score, 1 vs >=2 | 1.295 | 1.113 | 1.507 |
| Frailty, Quartile 1 vs Quartile 4 | 1.078 | 0.991 | 1.173 |
| Frailty, Quartile 2 vs Quartile 4 | 1.105 | 1.004 | 1.215 |
| Frailty, Quartile 3 vs Quartile 4 | 1.109 | 1.024 | 1.2 |
| Specimen type, Nasopharyngeal/oropharyngeal swab vs Saliva | 1.781 | 1.662 | 1.909 |
| Immunocompromised status, No vs Yes | 1.512 | 1.235 | 1.851 |
| History of COVID-19 diagnosis, No vs Yes | 1.53 | 1.43 | 1.637 |

### Supplementary Table 4. Full model for 2-dose VE against delta variant infection

| Odds Ratio Estimates | | | |
|---|---|---|---|
| Effect | Point Estimate | 95% Wald Confidence Interval | |
| Vaccination status, Vaccinated vs Unvaccinated | 0.364 | 0.331 | 0.401 |
| History of SARS-CoV-2 molecular test, No vs Yes | 1.222 | 1.108 | 1.347 |
| Preventive Care, No vs Yes | 1.064 | 0.96 | 1.178 |
| Number of outpatient and virtual visits, 0 vs >=11 | 1.541 | 1.243 | 1.909 |
| Number of outpatient and virtual visits, 1-4 vs >=11 | 1.425 | 1.248 | 1.628 |
| Number of outpatient and virtual visits, 5-10 vs >=11 | 1.188 | 1.053 | 1.341 |
| Obese, No vs Yes | 0.882 | 0.801 | 0.971 |
| Obese, Unknown vs Yes | 0.913 | 0.784 | 1.063 |
| Charlson comorbidity score, 0 vs >=2 | 1.154 | 0.958 | 1.389 |
| Charlson comorbidity score, 1 vs >=2 | 1.309 | 1.069 | 1.603 |
| Frailty, Quartile 1 vs Quartile 4 | 1.209 | 1.049 | 1.394 |
| Frailty, Quartile 2 vs Quartile 4 | 1.205 | 1.033 | 1.405 |
| Frailty, Quartile 3 vs Quartile 4 | 1.173 | 1.025 | 1.341 |
| Specimen type, Nasopharyngeal/oropharyngeal swab vs Saliva | 3.455 | 3.063 | 3.897 |
| Immunocompromised status, No vs Yes | 1.635 | 1.203 | 2.224 |
| History of COVID-19 diagnosis, No vs Yes | 10.112 | 8.142 | 12.558 |

Supplementary Table 5. Full model for 2-dose VE against delta variant infection by time since vaccination

| Odds Ratio Estimates | | | |
|---|---|---|---|
| Effect | Point Estimate | 95% Wald Confidence Interval | |
| Vaccination status, Vaccinated 14-90 days vs Unvaccinated | 0.198 | 0.123 | 0.318 |
| Vaccination status, Vaccinated 91-180 days vs Unvaccinated | 0.311 | 0.242 | 0.399 |
| Vaccination status, Vaccinated 181-270 days vs Unvaccinated | 0.363 | 0.328 | 0.402 |
| Vaccination status, Vaccinated 271-365 days vs Unvaccinated | 0.387 | 0.333 | 0.45 |
| History of SARS-CoV-2 molecular test, No vs Yes | 1.23 | 1.12 | 1.352 |
| Preventive Care, No vs Yes | 1.07 | 0.971 | 1.179 |
| Number of outpatient and virtual visits, 0 vs >=11 | 1.459 | 1.189 | 1.79 |
| Number of outpatient and virtual visits, 1-4 vs >=11 | 1.405 | 1.239 | 1.594 |
| Number of outpatient and virtual visits, 5-10 vs >=11 | 1.177 | 1.049 | 1.32 |
| Obese, No vs Yes | 0.906 | 0.827 | 0.993 |
| Obese, Unknown vs Yes | 0.952 | 0.824 | 1.1 |
| Charlson comorbidity score, 0 vs >=2 | 1.126 | 0.944 | 1.344 |
| Charlson comorbidity score, 1 vs >=2 | 1.235 | 1.02 | 1.497 |
| Frailty, Quartile 1 vs Quartile 4 | 1.177 | 1.027 | 1.348 |
| Frailty, Quartile 2 vs Quartile 4 | 1.15 | 0.993 | 1.332 |
| Frailty, Quartile 3 vs Quartile 4 | 1.125 | 0.99 | 1.279 |
| Specimen type, Nasopharyngeal/oropharyngeal swab vs Saliva | 3.575 | 3.189 | 4.008 |
| Immunocompromised status, No vs Yes | 1.731 | 1.293 | 2.317 |
| History of COVID-19 diagnosis, No vs Yes | 9.929 | 8.06 | 12.231 |
| Age group, 18-44 years old vs 75 and older | 0.735 | 0.533 | 1.013 |
| Age group, 45-64 years old vs 75 and older | 0.844 | 0.615 | 1.159 |
| Age group, 65-74 years old vs 75 and older | 0.865 | 0.616 | 1.214 |
| Sex, Female vs Male | 1.185 | 1.087 | 1.292 |
| Race/ethnicity, Hispanic vs Other/Unknown | 1.157 | 0.986 | 1.358 |
| Race/ethnicity, Non-Hispanic Asian vs Other/Unknown | 1.336 | 1.047 | 1.706 |
| Race/ethnicity, Non-Hispanic Black vs Other/Unknown | 1.039 | 0.829 | 1.303 |
| Race/ethnicity, Non-Hispanic White vs Other/Unknown | 1.067 | 0.907 | 1.254 |

### Supplementary Table 6. Full model for 2-dose VE against omicron variant infection

| Odds Ratio Estimates | | | |
|---|---|---|---|
| Effect | Point Estimate | 95% Wald Confidence Interval | |
| Vaccination status, Vaccinated vs Unvaccinated | 0.861 | 0.829 | 0.895 |
| History of SARS-CoV-2 molecular test, No vs Yes | 0.943 | 0.905 | 0.984 |
| Preventive Care, No vs Yes | 1.12 | 1.075 | 1.168 |
| Number of outpatient and virtual visits, 0 vs >=11 | 1.413 | 1.287 | 1.552 |
| Number of outpatient and virtual visits, 1-4 vs >=11 | 1.379 | 1.305 | 1.457 |
| Number of outpatient and virtual visits, 5-10 vs >=11 | 1.267 | 1.206 | 1.33 |
| Obese, No vs Yes | 0.942 | 0.905 | 0.98 |
| Obese, Unknown vs Yes | 0.985 | 0.925 | 1.05 |
| Charlson comorbidity score, 0 vs >=2 | 1.314 | 1.208 | 1.43 |
| Charlson comorbidity score, 1 vs >=2 | 1.274 | 1.162 | 1.396 |
| Frailty, Quartile 1 vs Quartile 4 | 1.091 | 1.031 | 1.155 |
| Frailty, Quartile 2 vs Quartile 4 | 1.099 | 1.031 | 1.171 |
| Frailty, Quartile 3 vs Quartile 4 | 1.101 | 1.044 | 1.162 |
| Specimen type, Nasopharyngeal/oropharyngeal swab vs Saliva | 1.892 | 1.796 | 1.993 |
| Immunocompromised status, No vs Yes | 1.34 | 1.178 | 1.524 |
| History of COVID-19 diagnosis, No vs Yes | 1.639 | 1.558 | 1.724 |

Supplementary Table 7. Full model for 2-dose VE against omicron variant infection by time since vaccination

| Odds Ratio Estimates | | | |
|---|---|---|---|
| **Effect** | **Point Estimate** | **95% Wald Confidence Interval** | |
| Vaccination status, Vaccinated 14-90 days vs Unvaccinated | 0.56 | 0.484 | 0.649 |
| Vaccination status, Vaccinated 91-180 days vs Unvaccinated | 0.765 | 0.7 | 0.836 |
| Vaccination status, Vaccinated 181-270 days vs Unvaccinated | 0.862 | 0.827 | 0.898 |
| Vaccination status, Vaccinated 271-365 days vs Unvaccinated | 0.941 | 0.89 | 0.996 |
| History of SARS-CoV-2 molecular test, No vs Yes | 0.939 | 0.901 | 0.979 |
| Preventive Care, No vs Yes | 1.129 | 1.083 | 1.177 |
| Number of outpatient and virtual visits, 0 vs >=11 | 1.396 | 1.271 | 1.532 |
| Number of outpatient and virtual visits, 1-4 vs >=11 | 1.368 | 1.296 | 1.445 |
| Number of outpatient and virtual visits, 5-10 vs >=11 | 1.26 | 1.2 | 1.322 |
| Obese, No vs Yes | 0.94 | 0.904 | 0.978 |
| Obese, Unknown vs Yes | 0.988 | 0.928 | 1.051 |
| Charlson comorbidity score, 0 vs >=2 | 1.312 | 1.206 | 1.428 |
| Charlson comorbidity score, 1 vs >=2 | 1.268 | 1.157 | 1.389 |
| Frailty, Quartile 1 vs Quartile 4 | 1.098 | 1.037 | 1.162 |
| Frailty, Quartile 2 vs Quartile 4 | 1.102 | 1.035 | 1.174 |
| Frailty, Quartile 3 vs Quartile 4 | 1.105 | 1.047 | 1.166 |
| Specimen type, Nasopharyngeal/oropharyngeal swab vs Saliva | 1.897 | 1.801 | 1.998 |
| Immunocompromised status, No vs Yes | 1.344 | 1.182 | 1.528 |
| History of COVID-19 diagnosis, No vs Yes | 1.638 | 1.557 | 1.723 |
| Age group, 18-44 years old vs 75 and older | 0.729 | 0.609 | 0.872 |
| Age group, 45-64 years old vs 75 and older | 0.818 | 0.684 | 0.978 |
| Age group, 65-74 years old vs 75 and older | 0.902 | 0.742 | 1.097 |
| Sex, Female vs Male | 1.106 | 1.066 | 1.148 |
| Race/ethnicity, Hispanic vs Other/Unknown | 1.067 | 0.999 | 1.14 |
| Race/ethnicity, Non-Hispanic Asian vs Other/Unknown | 1.049 | 0.961 | 1.144 |
| Race/ethnicity, Non-Hispanic Black vs Other/Unknown | 1.092 | 1.003 | 1.188 |
| Race/ethnicity, Non-Hispanic White vs Other/Unknown | 1.05 | 0.979 | 1.126 |

### Supplementary Table 8. Full model for 3-dose VE against delta variant infection

| Odds Ratio Estimates | | | |
|---|---|---|---|
| **Effect** | **Point Estimate** | **95% Wald Confidence Interval** | |
| Vaccination status, Vaccinated vs Unvaccinated | 0.055 | 0.043 | 0.071 |
| History of SARS-CoV-2 molecular test, No vs Yes | 1.391 | 1.226 | 1.578 |
| Preventive Care, No vs Yes | 1.176 | 1.031 | 1.342 |
| Number of outpatient and virtual visits, 0 vs >=11 | 1.434 | 1.105 | 1.861 |
| Number of outpatient and virtual visits, 1-4 vs >=11 | 1.407 | 1.187 | 1.667 |
| Number of outpatient and virtual visits, 5-10 vs >=11 | 1.138 | 0.97 | 1.336 |
| Obese, No vs Yes | 0.881 | 0.78 | 0.996 |
| Obese, Unknown vs Yes | 0.92 | 0.762 | 1.111 |
| Charlson comorbidity score, 0 vs >=2 | 0.951 | 0.737 | 1.227 |
| Charlson comorbidity score, 1 vs >=2 | 1.216 | 0.921 | 1.606 |
| Frailty, Quartile 1 vs Quartile 4 | 1.145 | 0.953 | 1.374 |
| Frailty, Quartile 2 vs Quartile 4 | 1.042 | 0.852 | 1.275 |
| Frailty, Quartile 3 vs Quartile 4 | 1.051 | 0.884 | 1.249 |
| Specimen type, Nasopharyngeal/oropharyngeal swab vs Saliva | 3.673 | 3.191 | 4.229 |
| Immunocompromised status, No vs Yes | 1.935 | 1.298 | 2.884 |
| History of COVID-19 diagnosis, No vs Yes | 7.863 | 6.188 | 9.992 |

Supplementary Table 9. Full model for 3-dose VE against delta variant infection by time since vaccination

| Odds Ratio Estimates | | | |
|---|---|---|---|
| Effect | Point Estimate | 95% Wald Confidence Interval | |
| Vaccination status, Vaccinated 14-60 days vs Unvaccinated | 0.063 | 0.051 | 0.078 |
| Vaccination status, Vaccinated 61-365 days vs Unvaccinated | 0.14 | 0.089 | 0.219 |
| History of SARS-CoV-2 molecular test, No vs Yes | 1.4 | 1.246 | 1.572 |
| Preventive Care, No vs Yes | 1.154 | 1.022 | 1.303 |
| Number of outpatient and virtual visits, 0 vs >=11 | 1.453 | 1.14 | 1.851 |
| Number of outpatient and virtual visits, 1-4 vs >=11 | 1.414 | 1.209 | 1.653 |
| Number of outpatient and virtual visits, 5-10 vs >=11 | 1.175 | 1.013 | 1.362 |
| Obese, No vs Yes | 0.869 | 0.777 | 0.973 |
| Obese, Unknown vs Yes | 0.947 | 0.797 | 1.125 |
| Charlson comorbidity score, 0 vs >=2 | 0.972 | 0.77 | 1.226 |
| Charlson comorbidity score, 1 vs >=2 | 1.214 | 0.943 | 1.561 |
| Frailty, Quartile 1 vs Quartile 4 | 1.151 | 0.973 | 1.362 |
| Frailty, Quartile 2 vs Quartile 4 | 1.07 | 0.889 | 1.289 |
| Frailty, Quartile 3 vs Quartile 4 | 1.06 | 0.905 | 1.242 |
| Specimen type, Nasopharyngeal/oropharyngeal swab vs Saliva | 3.804 | 3.35 | 4.319 |
| Immunocompromised status, No vs Yes | 1.982 | 1.376 | 2.855 |
| History of COVID-19 diagnosis, No vs Yes | 8.137 | 6.498 | 10.191 |
| Age group, 18-44 years old vs 75 and older | 0.317 | 0.203 | 0.494 |
| Age group, 45-64 years old vs 75 and older | 0.432 | 0.278 | 0.669 |
| Age group, 65-74 years old vs 75 and older | 0.763 | 0.479 | 1.216 |
| Sex, Female vs Male | 1.26 | 1.132 | 1.402 |
| Race/ethnicity, Hispanic vs Other/Unknown | 0.971 | 0.801 | 1.178 |
| Race/ethnicity, Non-Hispanic Asian vs Other/Unknown | 1.964 | 1.414 | 2.726 |
| Race/ethnicity, Non-Hispanic Black vs Other/Unknown | 0.906 | 0.694 | 1.184 |
| Race/ethnicity, Non-Hispanic White vs Other/Unknown | 1.058 | 0.87 | 1.287 |

### Supplementary Table 10. Full model for 3-dose VE against omicron variant infection

| Odds Ratio Estimates | | | |
|---|---|---|---|
| Effect | Point Estimate | 95% Wald Confidence Interval | |
| Vaccination status, Vaccinated vs Unvaccinated | 0.3 | 0.281 | 0.32 |
| History of SARS-CoV-2 molecular test, No vs Yes | 0.917 | 0.865 | 0.972 |
| Preventive Care, No vs Yes | 1.073 | 1.011 | 1.14 |
| Number of outpatient and virtual visits, 0 vs >=11 | 1.566 | 1.384 | 1.772 |
| Number of outpatient and virtual visits, 1-4 vs >=11 | 1.377 | 1.278 | 1.484 |
| Number of outpatient and virtual visits, 5-10 vs >=11 | 1.208 | 1.13 | 1.291 |
| Obese, No vs Yes | 1.019 | 0.965 | 1.076 |
| Obese, Unknown vs Yes | 0.984 | 0.901 | 1.074 |
| Charlson comorbidity score, 0 vs >=2 | 1.092 | 0.981 | 1.216 |
| Charlson comorbidity score, 1 vs >=2 | 1.003 | 0.894 | 1.125 |
| Frailty, Quartile 1 vs Quartile 4 | 1.053 | 0.973 | 1.141 |
| Frailty, Quartile 2 vs Quartile 4 | 1.053 | 0.966 | 1.148 |
| Frailty, Quartile 3 vs Quartile 4 | 1.091 | 1.012 | 1.175 |
| Specimen type, Nasopharyngeal/oropharyngeal swab vs Saliva | 1.661 | 1.558 | 1.771 |
| Immunocompromised status, No vs Yes | 1.105 | 0.95 | 1.285 |
| History of COVID-19 diagnosis, No vs Yes | 1.469 | 1.373 | 1.572 |

Supplementary Table 11. Full model for 3-dose VE against omicron variant infection by time since vaccination

| Odds Ratio Estimates | | | |
|---|---|---|---|
| Effect | Point Estimate | 95% Wald Confidence Interval | |
| Vaccination status, Vaccinated 14-60 days vs Unvaccinated | 0.284 | 0.266 | 0.303 |
| Vaccination status, Vaccinated 61-365 days vs Unvaccinated | 0.526 | 0.465 | 0.595 |
| History of SARS-CoV-2 molecular test, No vs Yes | 0.916 | 0.865 | 0.97 |
| Preventive Care, No vs Yes | 1.077 | 1.015 | 1.143 |
| Number of outpatient and virtual visits, 0 vs >=11 | 1.57 | 1.392 | 1.772 |
| Number of outpatient and virtual visits, 1-4 vs >=11 | 1.387 | 1.289 | 1.492 |
| Number of outpatient and virtual visits, 5-10 vs >=11 | 1.227 | 1.149 | 1.31 |
| Obese, No vs Yes | 1.013 | 0.961 | 1.068 |
| Obese, Unknown vs Yes | 0.975 | 0.894 | 1.062 |
| Charlson comorbidity score, 0 vs >=2 | 1.119 | 1.007 | 1.244 |
| Charlson comorbidity score, 1 vs >=2 | 1.021 | 0.911 | 1.143 |
| Frailty, Quartile 1 vs Quartile 4 | 1.063 | 0.983 | 1.149 |
| Frailty, Quartile 2 vs Quartile 4 | 1.061 | 0.974 | 1.155 |
| Frailty, Quartile 3 vs Quartile 4 | 1.089 | 1.013 | 1.172 |
| Specimen type, Nasopharyngeal/oropharyngeal swab vs Saliva | 1.685 | 1.582 | 1.794 |
| Immunocompromised status, No vs Yes | 1.217 | 1.048 | 1.413 |
| History of COVID-19 diagnosis, No vs Yes | 1.477 | 1.382 | 1.579 |
| Age group, 18-44 years old vs 75 and older | 0.42 | 0.352 | 0.502 |
| Age group, 45-64 years old vs 75 and older | 0.599 | 0.503 | 0.714 |
| Age group, 65-74 years old vs 75 and older | 0.887 | 0.736 | 1.069 |
| Sex, Female vs Male | 1.135 | 1.079 | 1.193 |
| Race/ethnicity, Hispanic vs Other/Unknown | 0.977 | 0.891 | 1.071 |
| Race/ethnicity, Non-Hispanic Asian vs Other/Unknown | 1.443 | 1.274 | 1.635 |
| Race/ethnicity, Non-Hispanic Black vs Other/Unknown | 0.939 | 0.839 | 1.052 |
| Race/ethnicity, Non-Hispanic White vs Other/Unknown | 1.015 | 0.921 | 1.117 |

Supplementary Table 12. Full model for 3-dose VE against delta variant infection by time since vaccination (excluding immunocompromised population)

| Odds Ratio Estimates | | | |
|---|---|---|---|
| Effect | Point Estimate | 95% Wald Confidence Interval | |
| Vaccination status, Vaccinated 14-60 days vs Unvaccinated | 0.058 | 0.047 | 0.073 |
| Vaccination status, Vaccinated 61-365 days vs Unvaccinated | 0.119 | 0.071 | 0.198 |
| History of SARS-CoV-2 molecular test, No vs Yes | 1.388 | 1.234 | 1.561 |
| Preventive Care, No vs Yes | 1.144 | 1.012 | 1.293 |
| Number of outpatient and virtual visits, 0 vs >=11 | 1.467 | 1.148 | 1.876 |
| Number of outpatient and virtual visits, 1-4 vs >=11 | 1.429 | 1.22 | 1.674 |
| Number of outpatient and virtual visits, 5-10 vs >=11 | 1.193 | 1.026 | 1.386 |
| Obese, No vs Yes | 0.861 | 0.768 | 0.965 |
| Obese, Unknown vs Yes | 0.933 | 0.784 | 1.109 |
| Charlson comorbidity score, 0 vs >=2 | 0.945 | 0.742 | 1.204 |
| Charlson comorbidity score, 1 vs >=2 | 1.159 | 0.891 | 1.507 |
| Frailty, Quartile 1 vs Quartile 4 | 1.157 | 0.977 | 1.37 |
| Frailty, Quartile 2 vs Quartile 4 | 1.093 | 0.904 | 1.32 |
| Frailty, Quartile 3 vs Quartile 4 | 1.096 | 0.933 | 1.287 |
| Specimen type, Nasopharyngeal/oropharyngeal swab vs Saliva | 3.79 | 3.335 | 4.307 |
| History of COVID-19 diagnosis, No vs Yes | 8.248 | 6.569 | 10.356 |
| Age group, 18-44 years old vs 75 and older | 0.308 | 0.195 | 0.489 |
| Age group, 45-64 years old vs 75 and older | 0.419 | 0.266 | 0.661 |
| Age group, 65-74 years old vs 75 and older | 0.786 | 0.485 | 1.275 |
| Sex, Female vs Male | 1.264 | 1.134 | 1.408 |
| Race/ethnicity, Hispanic vs Other/Unknown | 0.97 | 0.799 | 1.179 |
| Race/ethnicity, Non-Hispanic Asian vs Other/Unknown | 1.96 | 1.404 | 2.735 |
| Race/ethnicity, Non-Hispanic Black vs Other/Unknown | 0.881 | 0.673 | 1.154 |
| Race/ethnicity, Non-Hispanic White vs Other/Unknown | 1.07 | 0.879 | 1.304 |

Supplementary Table 13. Full model for 3-dose VE against omicron variant infection by time since vaccination (excluding immunocompromised population)

| Odds Ratio Estimates | | | |
|---|---|---|---|
| **Effect** | **Point Estimate** | **95% Wald Confidence Interval** | |
| Vaccination status, Vaccinated 14-60 days vs Unvaccinated | 0.279 | 0.261 | 0.298 |
| Vaccination status, Vaccinated 61-365 days vs Unvaccinated | 0.488 | 0.427 | 0.558 |
| History of SARS-CoV-2 molecular test, No vs Yes | 0.91 | 0.859 | 0.964 |
| Preventive Care, No vs Yes | 1.067 | 1.004 | 1.133 |
| Number of outpatient and virtual visits, 0 vs >=11 | 1.566 | 1.385 | 1.769 |
| Number of outpatient and virtual visits, 1-4 vs >=11 | 1.381 | 1.282 | 1.487 |
| Number of outpatient and virtual visits, 5-10 vs >=11 | 1.217 | 1.138 | 1.301 |
| Obese, No vs Yes | 1.015 | 0.962 | 1.071 |
| Obese, Unknown vs Yes | 0.978 | 0.896 | 1.066 |
| Charlson comorbidity score, 0 vs >=2 | 1.136 | 1.016 | 1.269 |
| Charlson comorbidity score, 1 vs >=2 | 1.042 | 0.925 | 1.174 |
| Frailty, Quartile 1 vs Quartile 4 | 1.065 | 0.985 | 1.153 |
| Frailty, Quartile 2 vs Quartile 4 | 1.066 | 0.978 | 1.163 |
| Frailty, Quartile 3 vs Quartile 4 | 1.089 | 1.011 | 1.172 |
| Specimen type, Nasopharyngeal/oropharyngeal swab vs Saliva | 1.704 | 1.599 | 1.817 |
| History of COVID-19 diagnosis, No vs Yes | 1.478 | 1.382 | 1.581 |
| Age group, 18-44 years old vs 75 and older | 0.399 | 0.332 | 0.481 |
| Age group, 45-64 years old vs 75 and older | 0.568 | 0.473 | 0.683 |
| Age group, 65-74 years old vs 75 and older | 0.854 | 0.704 | 1.037 |
| Sex, Female vs Male | 1.139 | 1.083 | 1.199 |
| Race/ethnicity, Hispanic vs Other/Unknown | 0.973 | 0.887 | 1.069 |
| Race/ethnicity, Non-Hispanic Asian vs Other/Unknown | 1.461 | 1.287 | 1.658 |
| Race/ethnicity, Non-Hispanic Black vs Other/Unknown | 0.937 | 0.836 | 1.051 |
| Race/ethnicity, Non-Hispanic White vs Other/Unknown | 1.015 | 0.92 | 1.118 |

### Supplementary Table 14. Full model for 1-dose VE against delta variant hospitalization

**Odds Ratio Estimates**

| Effect | Point Estimate | 95% Wald Confidence Interval | |
|---|---|---|---|
| Vaccination status, Vaccinated vs Unvaccinated | 0.288 | 0.026 | 3.194 |
| History of SARS-CoV-2 molecular test, No vs Yes | 1.59 | 0.795 | 3.182 |
| Preventive Care, No vs Yes | 2.393 | 1.041 | 5.502 |
| Obese, No vs Yes | 0.166 | 0.066 | 0.413 |
| Obese, Unknown vs Yes | 0.268 | 0.082 | 0.882 |
| Charlson comorbidity score, 0 vs >=2 | 0.592 | 0.237 | 1.481 |
| Charlson comorbidity score, 1 vs >=2 | 0.902 | 0.282 | 2.88 |
| History of COVID-19 diagnosis, No vs Yes | 10.386 | 2.246 | 48.021 |

### Supplementary Table 15. Full model for 2-dose VE against delta variant hospitalization

**Odds Ratio Estimates**

| Effect | Point Estimate | 95% Wald Confidence Interval | |
|---|---|---|---|
| Vaccination status, Vaccinated vs Unvaccinated | 0.01 | 0.001 | 0.067 |
| History of SARS-CoV-2 molecular test, No vs Yes | 0.734 | 0.267 | 2.02 |
| Preventive Care, No vs Yes | 4.08 | 1.353 | 12.307 |
| Obese, No vs Yes | 0.092 | 0.024 | 0.349 |
| Obese, Unknown vs Yes | 0.546 | 0.132 | 2.265 |
| Charlson comorbidity score, 0 vs >=2 | 0.219 | 0.039 | 1.224 |
| Charlson comorbidity score, 1 vs >=2 | 1.259 | 0.197 | 8.031 |
| History of COVID-19 diagnosis, No vs Yes | 18.718 | 2.917 | 120.099 |

### Supplementary Table 16. Full model for 2-dose VE against omicron variant hospitalization

| Odds Ratio Estimates | | | |
|---|---|---|---|
| **Effect** | **Point Estimate** | **95% Wald Confidence Interval** | |
| Vaccination status, Vaccinated vs Unvaccinated | 0.155 | 0.031 | 0.77 |
| History of SARS-CoV-2 molecular test, No vs Yes | 4.677 | 0.635 | 34.442 |
| Preventive Care, No vs Yes | 0.717 | 0.146 | 3.515 |
| Charlson comorbidity score, 0 vs >=2 | 0.902 | 0.083 | 9.842 |
| Charlson comorbidity score, 1 vs >=2 | 3.359 | 0.237 | 47.543 |
| Immunocompromised status, No vs Yes | 0.175 | 0.006 | 5.289 |
| History of COVID-19 diagnosis, No vs Yes | 2.364 | 0.172 | 32.54 |

### Supplementary Table 17. Full model for 3-dose VE against delta variant hospitalization

| Odds Ratio Estimates | | | |
|---|---|---|---|
| **Effect** | **Point Estimate** | **95% Wald Confidence Interval** | |
| Vaccination status, Vaccinated vs Unvaccinated | 0.003 | <0.001 | 0.035 |
| History of SARS-CoV-2 molecular test, No vs Yes | 1.738 | 0.79 | 3.825 |
| Preventive Care, No vs Yes | 1.715 | 0.759 | 3.875 |
| Obese, No vs Yes | 0.322 | 0.136 | 0.762 |
| Obese, Unknown vs Yes | 0.418 | 0.14 | 1.249 |
| Charlson comorbidity score, 0 vs >=2 | 0.313 | 0.105 | 0.931 |
| Charlson comorbidity score, 1 vs >=2 | 0.479 | 0.131 | 1.753 |
| History of COVID-19 diagnosis, No vs Yes | 8.736 | 1.711 | 44.601 |
| Age group, 18-44 years old vs 75 and older | 0.202 | 0.031 | 1.312 |
| Age group, 45-64 years old vs 75 and older | 0.23 | 0.039 | 1.363 |
| Age group, 65-74 years old vs 75 and older | 1.484 | 0.177 | 12.452 |
| Sex, Female vs Male | 1.157 | 0.53 | 2.524 |
| Race/ethnicity, Hispanic vs Other/Unknown | 0.817 | 0.202 | 3.304 |
| Race/ethnicity, Non-Hispanic Asian vs Other/Unknown | 1.146 | 0.138 | 9.498 |
| Race/ethnicity, Non-Hispanic Black vs Other/Unknown | 1.428 | 0.254 | 8.024 |
| Race/ethnicity, Non-Hispanic White vs Other/Unknown | 0.858 | 0.202 | 3.638 |

### Supplementary Table 18. Full model for 3-dose VE against omicron variant hospitalization

| Odds Ratio Estimates | | | |
|---|---|---|---|
| Effect | Point Estimate | 95% Wald Confidence Interval | |
| Vaccination status, Vaccinated vs Unvaccinated | 0.008 | <0.001 | 0.237 |
| History of SARS-CoV-2 molecular test, No vs Yes | 0.473 | 0.059 | 3.765 |
| Preventive Care, No vs Yes | 1.008 | 0.075 | 13.531 |
| Charlson comorbidity score, 0 vs >=2 | 0.033 | 0.002 | 0.725 |
| Charlson comorbidity score, 1 vs >=2 | 0.765 | 0.02 | 29.284 |
| Immunocompromised status, No vs Yes | 0.261 | 0.007 | 9.422 |
| Age group, 18-44 years old vs 75 and older | 0.862 | 0.052 | 14.438 |
| Age group, 45-64 years old vs 75 and older | 2.605 | 0.099 | 68.699 |
| Age group, 65-74 years old vs 75 and older | 1.62 | 0.078 | 33.492 |
| Sex, Female vs Male | 0.216 | 0.02 | 2.356 |
| Race/ethnicity, Hispanic vs Non-Hispanic White | 3.084 | 0.2 | 47.486 |
| Race/ethnicity, Non-Hispanic Asian vs Non-Hispanic White | 5.291 | 0.124 | 224.949 |
| Race/ethnicity, Non-Hispanic Black vs Non-Hispanic White | 0.435 | 0.024 | 7.917 |

Supplementary Table 19. Full model for 3-dose VE against delta variant infection (subgroup of age<65)

| Odds Ratio Estimates | | | |
|---|---|---|---|
| **Effect** | **Point Estimate** | **95% Wald Confidence Interval** | |
| Vaccination status, Vaccinated vs Unvaccinated | 0.057 | 0.043 | 0.075 |
| History of SARS-CoV-2 molecular test, No vs Yes | 1.416 | 1.243 | 1.614 |
| Preventive Care, No vs Yes | 1.18 | 1.032 | 1.349 |
| Number of outpatient and virtual visits, 0 vs >=11 | 1.42 | 1.084 | 1.86 |
| Number of outpatient and virtual visits, 1-4 vs >=11 | 1.412 | 1.186 | 1.682 |
| Number of outpatient and virtual visits, 5-10 vs >=11 | 1.158 | 0.981 | 1.368 |
| Obese, No vs Yes | 0.895 | 0.788 | 1.016 |
| Obese, Unknown vs Yes | 0.927 | 0.765 | 1.123 |
| Charlson comorbidity score, 0 vs >=2 | 0.939 | 0.707 | 1.246 |
| Charlson comorbidity score, 1 vs >=2 | 1.186 | 0.866 | 1.623 |
| Frailty, Quartile 1 vs Quartile 4 | 1.176 | 0.981 | 1.41 |
| Frailty, Quartile 2 vs Quartile 4 | 1.1 | 0.893 | 1.356 |
| Frailty, Quartile 3 vs Quartile 4 | 1.096 | 0.921 | 1.306 |
| Specimen type, Nasopharyngeal/oropharyngeal swab vs Saliva | 3.833 | 3.315 | 4.432 |
| Immunocompromised status, No vs Yes | 1.754 | 1.14 | 2.699 |
| History of COVID-19 diagnosis, No vs Yes | 7.724 | 6.063 | 9.84 |

Supplementary Table 20. Full model for 3-dose VE against delta variant infection (subgroup of age≥65)

| Odds Ratio Estimates | | | |
|---|---|---|---|
| Effect | Point Estimate | 95% Wald Confidence Interval | |
| Vaccination status, Vaccinated vs Unvaccinated | 0.04 | 0.021 | 0.077 |
| History of SARS-CoV-2 molecular test, No vs Yes | 1.127 | 0.674 | 1.884 |
| Preventive Care, No vs Yes | 1.119 | 0.522 | 2.399 |
| Number of outpatient and virtual visits, 0 vs >=11 | 0.945 | 0.156 | 5.734 |
| Number of outpatient and virtual visits, 1-4 vs >=11 | 1.002 | 0.441 | 2.276 |
| Number of outpatient and virtual visits, 5-10 vs >=11 | 0.682 | 0.365 | 1.276 |
| Obese, No vs Yes | 0.633 | 0.378 | 1.062 |
| Obese, Unknown vs Yes | 0.671 | 0.175 | 2.579 |
| Charlson comorbidity score, 0 vs >=2 | 0.814 | 0.401 | 1.653 |
| Charlson comorbidity score, 1 vs >=2 | 1.259 | 0.639 | 2.482 |
| Frailty, Quartile 1 vs Quartile 4 | 2.186 | 0.942 | 5.076 |
| Frailty, Quartile 2 vs Quartile 4 | 2.095 | 0.962 | 4.564 |
| Frailty, Quartile 3 vs Quartile 4 | 2.14 | 1.011 | 4.527 |
| Specimen type, Nasopharyngeal/oropharyngeal swab vs Saliva | 1.503 | 0.771 | 2.93 |
| Immunocompromised status, No vs Yes | 4.096 | 1.381 | 12.154 |
| History of COVID-19 diagnosis, No vs Yes | 10.522 | 2.028 | 54.586 |

Supplementary Table 21. Full model for 3-dose VE against delta variant infection (subgroup of female)

| Odds Ratio Estimates | | | |
|---|---|---|---|
| Effect | Point Estimate | 95% Wald Confidence Interval | |
| Vaccination status, Vaccinated vs Unvaccinated | 0.056 | 0.04 | 0.078 |
| History of SARS-CoV-2 molecular test, No vs Yes | 1.411 | 1.172 | 1.7 |
| Preventive Care, No vs Yes | 1.123 | 0.94 | 1.343 |
| Number of outpatient and virtual visits, 0 vs >=11 | 1.863 | 1.27 | 2.732 |
| Number of outpatient and virtual visits, 1-4 vs >=11 | 1.464 | 1.173 | 1.828 |
| Number of outpatient and virtual visits, 5-10 vs >=11 | 1.33 | 1.086 | 1.629 |
| Obese, No vs Yes | 0.895 | 0.76 | 1.052 |
| Obese, Unknown vs Yes | 0.765 | 0.568 | 1.031 |
| Charlson comorbidity score, 0 vs >=2 | 1.004 | 0.708 | 1.425 |
| Charlson comorbidity score, 1 vs >=2 | 1.16 | 0.796 | 1.69 |
| Frailty, Quartile 1 vs Quartile 4 | 1.075 | 0.837 | 1.381 |
| Frailty, Quartile 2 vs Quartile 4 | 0.95 | 0.73 | 1.236 |
| Frailty, Quartile 3 vs Quartile 4 | 1.014 | 0.8 | 1.286 |
| Specimen type, Nasopharyngeal/oropharyngeal swab vs Saliva | 3.769 | 3.115 | 4.56 |
| Immunocompromised status, No vs Yes | 1.606 | 0.97 | 2.662 |
| History of COVID-19 diagnosis, No vs Yes | 7.264 | 5.311 | 9.937 |

Supplementary Table 22. Full model for 3-dose VE against delta variant infection (subgroup of male)

| Odds Ratio Estimates | | | |
|---|---|---|---|
| **Effect** | **Point Estimate** | **95% Wald Confidence Interval** | |
| Vaccination status, Vaccinated vs Unvaccinated | 0.054 | 0.037 | 0.08 |
| History of SARS-CoV-2 molecular test, No vs Yes | 1.392 | 1.171 | 1.654 |
| Preventive Care, No vs Yes | 1.249 | 1.024 | 1.524 |
| Number of outpatient and virtual visits, 0 vs >=11 | 1.141 | 0.798 | 1.633 |
| Number of outpatient and virtual visits, 1-4 vs >=11 | 1.306 | 0.992 | 1.72 |
| Number of outpatient and virtual visits, 5-10 vs >=11 | 0.914 | 0.699 | 1.195 |
| Obese, No vs Yes | 0.871 | 0.721 | 1.052 |
| Obese, Unknown vs Yes | 1.027 | 0.8 | 1.32 |
| Charlson comorbidity score, 0 vs >=2 | 0.886 | 0.608 | 1.292 |
| Charlson comorbidity score, 1 vs >=2 | 1.294 | 0.853 | 1.964 |
| Frailty, Quartile 1 vs Quartile 4 | 1.209 | 0.928 | 1.575 |
| Frailty, Quartile 2 vs Quartile 4 | 1.052 | 0.682 | 1.623 |
| Frailty, Quartile 3 vs Quartile 4 | 1.114 | 0.871 | 1.423 |
| Specimen type, Nasopharyngeal/oropharyngeal swab vs Saliva | 3.54 | 2.868 | 4.368 |
| Immunocompromised status, No vs Yes | 2.598 | 1.356 | 4.979 |
| History of COVID-19 diagnosis, No vs Yes | 8.964 | 6.166 | 13.032 |

Supplementary Table 23. Full model for 3-dose VE against delta variant infection (subgroup of Hispanic)

**Odds Ratio Estimates**

| Effect | Point Estimate | 95% Wald Confidence Interval | |
|---|---|---|---|
| Vaccination status, Vaccinated vs Unvaccinated | 0.069 | 0.045 | 0.106 |
| History of SARS-CoV-2 molecular test, No vs Yes | 1.34 | 1.1 | 1.633 |
| Preventive Care, No vs Yes | 1.215 | 0.996 | 1.481 |
| Number of outpatient and virtual visits, 0 vs >=11 | 1.496 | 1.009 | 2.217 |
| Number of outpatient and virtual visits, 1-4 vs >=11 | 1.47 | 1.135 | 1.904 |
| Number of outpatient and virtual visits, 5-10 vs >=11 | 1.256 | 0.977 | 1.614 |
| Obese, No vs Yes | 0.972 | 0.809 | 1.169 |
| Obese, Unknown vs Yes | 1.092 | 0.826 | 1.444 |
| Charlson comorbidity score, 0 vs >=2 | 0.728 | 0.481 | 1.101 |
| Charlson comorbidity score, 1 vs >=2 | 0.875 | 0.56 | 1.366 |
| Frailty, Quartile 1 vs Quartile 4 | 1.096 | 0.832 | 1.443 |
| Frailty, Quartile 2 vs Quartile 4 | 1.038 | 0.767 | 1.404 |
| Frailty, Quartile 3 vs Quartile 4 | 1.161 | 0.894 | 1.507 |
| Specimen type, Nasopharyngeal/oropharyngeal swab vs Saliva | 4.264 | 3.377 | 5.384 |
| Immunocompromised status, No vs Yes | 2.077 | 1.083 | 3.982 |
| History of COVID-19 diagnosis, No vs Yes | 9.321 | 6.63 | 13.104 |

Supplementary Table 24. Full model for 3-dose VE against delta variant infection (subgroup of Non-Hispanic and others)

| Odds Ratio Estimates | | | |
|---|---|---|---|
| **Effect** | **Point Estimate** | **95% Wald Confidence Interval** | |
| Vaccination status, Vaccinated vs Unvaccinated | 0.049 | 0.036 | 0.068 |
| History of SARS-CoV-2 molecular test, No vs Yes | 1.452 | 1.231 | 1.712 |
| Preventive Care, No vs Yes | 1.143 | 0.958 | 1.365 |
| Number of outpatient and virtual visits, 0 vs >=11 | 1.372 | 0.967 | 1.946 |
| Number of outpatient and virtual visits, 1-4 vs >=11 | 1.323 | 1.055 | 1.658 |
| Number of outpatient and virtual visits, 5-10 vs >=11 | 1.029 | 0.835 | 1.269 |
| Obese, No vs Yes | 0.805 | 0.682 | 0.951 |
| Obese, Unknown vs Yes | 0.792 | 0.611 | 1.026 |
| Charlson comorbidity score, 0 vs >=2 | 1.117 | 0.806 | 1.549 |
| Charlson comorbidity score, 1 vs >=2 | 1.515 | 1.058 | 2.169 |
| Frailty, Quartile 1 vs Quartile 4 | 1.213 | 0.949 | 1.55 |
| Frailty, Quartile 2 vs Quartile 4 | 1.075 | 0.821 | 1.408 |
| Frailty, Quartile 3 vs Quartile 4 | 0.998 | 0.792 | 1.258 |
| Specimen type, Nasopharyngeal/oropharyngeal swab vs Saliva | 3.349 | 2.803 | 4.003 |
| Immunocompromised status, No vs Yes | 1.863 | 1.12 | 3.1 |
| History of COVID-19 diagnosis, No vs Yes | 6.581 | 4.69 | 9.236 |

Supplementary Table 25. Full model for 3-dose VE against delta variant infection (subgroup of immunocompromised population)

| Odds Ratio Estimates | | | |
|---|---|---|---|
| Effect | Point Estimate | 95% Wald Confidence Interval | |
| Vaccination status, Vaccinated vs Unvaccinated | 0.294 | 0.125 | 0.69 |
| History of SARS-CoV-2 molecular test, No vs Yes | 1.719 | 0.783 | 3.774 |
| Preventive Care, No vs Yes | 1.703 | 0.688 | 4.213 |
| Obese, No vs Yes | 1.364 | 0.619 | 3.003 |
| Obese, Unknown vs Yes | 3.835 | 0.448 | 32.845 |
| Charlson comorbidity score, 0 vs >=2 | 0.909 | 0.35 | 2.364 |
| Charlson comorbidity score, 1 vs >=2 | 2.397 | 0.912 | 6.297 |
| Frailty, Quartile 1 vs Quartile 4 | 2.594 | 0.884 | 7.615 |
| Frailty, Quartile 2 vs Quartile 4 | 0.856 | 0.276 | 2.658 |
| Frailty, Quartile 3 vs Quartile 4 | 1.325 | 0.457 | 3.839 |
| Specimen type, Nasopharyngeal/oropharyngeal swab vs Saliva | 12.207 | 2.606 | 57.185 |
| History of COVID-19 diagnosis, No vs Yes | 7.615 | 1.512 | 38.353 |
| Age group, 18-44 years old vs 75 and older | 0.268 | 0.035 | 2.052 |
| Age group, 45-64 years old vs 75 and older | 0.355 | 0.05 | 2.514 |
| Age group, 65-74 years old vs 75 and older | 0.323 | 0.043 | 2.42 |
| Sex, Female vs Male | 1.429 | 0.667 | 3.063 |
| Race/ethnicity, Hispanic vs Other/Unknown | 1.22 | 0.187 | 7.958 |
| Race/ethnicity, Non-Hispanic Asian vs Other/Unknown | 4.203 | 0.408 | 43.27 |
| Race/ethnicity, Non-Hispanic Black vs Other/Unknown | 3.04 | 0.358 | 25.813 |
| Race/ethnicity, Non-Hispanic White vs Other/Unknown | 0.778 | 0.12 | 5.06 |

Supplementary Table 26. Full model for 3-dose VE against delta variant infection (subgroup of non-immunocompromised)

**Odds Ratio Estimates**

| Effect | Point Estimate | 95% Wald Confidence Interval | |
|---|---|---|---|
| Vaccination status, Vaccinated vs Unvaccinated | 0.063 | 0.051 | 0.078 |
| History of SARS-CoV-2 molecular test, No vs Yes | 1.387 | 1.233 | 1.56 |
| Preventive Care, No vs Yes | 1.144 | 1.011 | 1.293 |
| Number of outpatient and virtual visits, 0 vs >=11 | 1.472 | 1.152 | 1.882 |
| Number of outpatient and virtual visits, 1-4 vs >=11 | 1.43 | 1.221 | 1.676 |
| Number of outpatient and virtual visits, 5-10 vs >=11 | 1.192 | 1.026 | 1.386 |
| Obese, No vs Yes | 0.861 | 0.768 | 0.966 |
| Obese, Unknown vs Yes | 0.933 | 0.785 | 1.11 |
| Charlson comorbidity score, 0 vs >=2 | 0.936 | 0.735 | 1.193 |
| Charlson comorbidity score, 1 vs >=2 | 1.153 | 0.886 | 1.5 |
| Frailty, Quartile 1 vs Quartile 4 | 1.159 | 0.979 | 1.373 |
| Frailty, Quartile 2 vs Quartile 4 | 1.092 | 0.904 | 1.319 |
| Frailty, Quartile 3 vs Quartile 4 | 1.098 | 0.935 | 1.29 |
| Specimen type, Nasopharyngeal/oropharyngeal swab vs Saliva | 3.789 | 3.334 | 4.306 |
| History of COVID-19 diagnosis, No vs Yes | 8.245 | 6.566 | 10.352 |
| Age group, 18-44 years old vs 75 and older | 0.292 | 0.184 | 0.464 |
| Age group, 45-64 years old vs 75 and older | 0.397 | 0.251 | 0.626 |
| Age group, 65-74 years old vs 75 and older | 0.752 | 0.462 | 1.222 |
| Sex, Female vs Male | 1.262 | 1.133 | 1.406 |
| Race/ethnicity, Hispanic vs Other/Unknown | 0.97 | 0.798 | 1.178 |
| Race/ethnicity, Non-Hispanic Asian vs Other/Unknown | 1.969 | 1.411 | 2.747 |
| Race/ethnicity, Non-Hispanic Black vs Other/Unknown | 0.881 | 0.673 | 1.153 |
| Race/ethnicity, Non-Hispanic White vs Other/Unknown | 1.073 | 0.881 | 1.307 |

Supplementary Table 27. Full model for 3-dose VE against omicron variant infection (subgroup of age<65)

| Odds Ratio Estimates | | | |
|---|---|---|---|
| **Effect** | **Point Estimate** | **95% Wald Confidence Interval** | |
| Vaccination status, Vaccinated vs Unvaccinated | 0.291 | 0.271 | 0.311 |
| History of SARS-CoV-2 molecular test, No vs Yes | 0.9 | 0.847 | 0.957 |
| Preventive Care, No vs Yes | 1.08 | 1.016 | 1.149 |
| Number of outpatient and virtual visits, 0 vs >=11 | 1.595 | 1.404 | 1.813 |
| Number of outpatient and virtual visits, 1-4 vs >=11 | 1.417 | 1.311 | 1.531 |
| Number of outpatient and virtual visits, 5-10 vs >=11 | 1.267 | 1.181 | 1.36 |
| Obese, No vs Yes | 1.033 | 0.976 | 1.093 |
| Obese, Unknown vs Yes | 0.991 | 0.906 | 1.084 |
| Charlson comorbidity score, 0 vs >=2 | 1.127 | 0.992 | 1.281 |
| Charlson comorbidity score, 1 vs >=2 | 1.038 | 0.903 | 1.193 |
| Frailty, Quartile 1 vs Quartile 4 | 1.037 | 0.956 | 1.124 |
| Frailty, Quartile 2 vs Quartile 4 | 1.056 | 0.965 | 1.155 |
| Frailty, Quartile 3 vs Quartile 4 | 1.051 | 0.973 | 1.135 |
| Specimen type, Nasopharyngeal/oropharyngeal swab vs Saliva | 1.701 | 1.59 | 1.819 |
| Immunocompromised status, No vs Yes | 1.063 | 0.899 | 1.258 |
| History of COVID-19 diagnosis, No vs Yes | 1.491 | 1.391 | 1.599 |

Supplementary Table 28. Full model for 3-dose VE against omicron variant infection (subgroup of age≥65)

**Odds Ratio Estimates**

| Effect | Point Estimate | 95% Wald Confidence Interval | |
|---|---|---|---|
| Vaccination status, Vaccinated vs Unvaccinated | 0.357 | 0.283 | 0.45 |
| History of SARS-CoV-2 molecular test, No vs Yes | 1.117 | 0.931 | 1.341 |
| Preventive Care, No vs Yes | 0.793 | 0.573 | 1.098 |
| Number of outpatient and virtual visits, 0 vs >=11 | 0.765 | 0.327 | 1.787 |
| Number of outpatient and virtual visits, 1-4 vs >=11 | 1.089 | 0.783 | 1.515 |
| Number of outpatient and virtual visits, 5-10 vs >=11 | 0.849 | 0.689 | 1.045 |
| Obese, No vs Yes | 0.869 | 0.727 | 1.04 |
| Obese, Unknown vs Yes | 0.732 | 0.432 | 1.238 |
| Charlson comorbidity score, 0 vs >=2 | 1.147 | 0.911 | 1.444 |
| Charlson comorbidity score, 1 vs >=2 | 1.034 | 0.827 | 1.294 |
| Frailty, Quartile 1 vs Quartile 4 | 1.061 | 0.794 | 1.417 |
| Frailty, Quartile 2 vs Quartile 4 | 1.076 | 0.834 | 1.389 |
| Frailty, Quartile 3 vs Quartile 4 | 1.04 | 0.819 | 1.319 |
| Specimen type, Nasopharyngeal/oropharyngeal swab vs Saliva | 1.316 | 1.055 | 1.641 |
| Immunocompromised status, No vs Yes | 1.232 | 0.868 | 1.75 |
| History of COVID-19 diagnosis, No vs Yes | 1.136 | 0.827 | 1.56 |

Supplementary Table 29. Full model for 3-dose VE against omicron variant infection (subgroup of female)

| Odds Ratio Estimates | | | |
|---|---|---|---|
| Effect | Point Estimate | 95% Wald Confidence Interval | |
| Vaccination status, Vaccinated vs Unvaccinated | 0.3 | 0.276 | 0.326 |
| History of SARS-CoV-2 molecular test, No vs Yes | 0.871 | 0.805 | 0.944 |
| Preventive Care, No vs Yes | 1.006 | 0.928 | 1.09 |
| Number of outpatient and virtual visits, 0 vs >=11 | 1.556 | 1.294 | 1.87 |
| Number of outpatient and virtual visits, 1-4 vs >=11 | 1.5 | 1.363 | 1.651 |
| Number of outpatient and virtual visits, 5-10 vs >=11 | 1.251 | 1.151 | 1.359 |
| Obese, No vs Yes | 1.043 | 0.973 | 1.119 |
| Obese, Unknown vs Yes | 0.996 | 0.874 | 1.135 |
| Charlson comorbidity score, 0 vs >=2 | 1.119 | 0.967 | 1.295 |
| Charlson comorbidity score, 1 vs >=2 | 1.036 | 0.887 | 1.21 |
| Frailty, Quartile 1 vs Quartile 4 | 0.991 | 0.89 | 1.102 |
| Frailty, Quartile 2 vs Quartile 4 | 1.012 | 0.908 | 1.128 |
| Frailty, Quartile 3 vs Quartile 4 | 1.096 | 0.994 | 1.209 |
| Specimen type, Nasopharyngeal/oropharyngeal swab vs Saliva | 1.578 | 1.45 | 1.716 |
| Immunocompromised status, No vs Yes | 1.204 | 0.986 | 1.472 |
| History of COVID-19 diagnosis, No vs Yes | 1.462 | 1.338 | 1.597 |

Supplementary Table 30. Full model for 3-dose VE against omicron variant infection (subgroup of male)

| Odds Ratio Estimates | | | |
|---|---|---|---|
| Effect | Point Estimate | 95% Wald Confidence Interval | |
| Vaccination status, Vaccinated vs Unvaccinated | 0.3 | 0.271 | 0.334 |
| History of SARS-CoV-2 molecular test, No vs Yes | 0.975 | 0.895 | 1.061 |
| Preventive Care, No vs Yes | 1.165 | 1.061 | 1.279 |
| Number of outpatient and virtual visits, 0 vs >=11 | 1.43 | 1.191 | 1.716 |
| Number of outpatient and virtual visits, 1-4 vs >=11 | 1.245 | 1.101 | 1.407 |
| Number of outpatient and virtual visits, 5-10 vs >=11 | 1.156 | 1.032 | 1.295 |
| Obese, No vs Yes | 0.978 | 0.898 | 1.066 |
| Obese, Unknown vs Yes | 0.946 | 0.839 | 1.068 |
| Charlson comorbidity score, 0 vs >=2 | 1.093 | 0.932 | 1.282 |
| Charlson comorbidity score, 1 vs >=2 | 0.983 | 0.827 | 1.169 |
| Frailty, Quartile 1 vs Quartile 4 | 1.113 | 0.986 | 1.256 |
| Frailty, Quartile 2 vs Quartile 4 | 1.111 | 0.966 | 1.278 |
| Frailty, Quartile 3 vs Quartile 4 | 1.017 | 0.906 | 1.142 |
| Specimen type, Nasopharyngeal/oropharyngeal swab vs Saliva | 1.78 | 1.612 | 1.966 |
| Immunocompromised status, No vs Yes | 0.978 | 0.776 | 1.233 |
| History of COVID-19 diagnosis, No vs Yes | 1.476 | 1.328 | 1.641 |

Supplementary Table 31. Full model for 3-dose VE against omicron variant infection (subgroup of Hispanic)

| Odds Ratio Estimates | | | |
|---|---|---|---|
| Effect | Point Estimate | 95% Wald Confidence Interval | |
| Vaccination status, Vaccinated vs Unvaccinated | 0.32 | 0.29 | 0.354 |
| History of SARS-CoV-2 molecular test, No vs Yes | 0.844 | 0.772 | 0.922 |
| Preventive Care, No vs Yes | 1.078 | 0.989 | 1.176 |
| Number of outpatient and virtual visits, 0 vs >=11 | 1.615 | 1.35 | 1.932 |
| Number of outpatient and virtual visits, 1-4 vs >=11 | 1.405 | 1.26 | 1.567 |
| Number of outpatient and virtual visits, 5-10 vs >=11 | 1.228 | 1.112 | 1.356 |
| Obese, No vs Yes | 1.009 | 0.933 | 1.09 |
| Obese, Unknown vs Yes | 1.045 | 0.919 | 1.188 |
| Charlson comorbidity score, 0 vs >=2 | 1.112 | 0.943 | 1.31 |
| Charlson comorbidity score, 1 vs >=2 | 1.081 | 0.908 | 1.287 |
| Frailty, Quartile 1 vs Quartile 4 | 1.027 | 0.914 | 1.153 |
| Frailty, Quartile 2 vs Quartile 4 | 1.031 | 0.909 | 1.169 |
| Frailty, Quartile 3 vs Quartile 4 | 1.134 | 1.016 | 1.265 |
| Specimen type, Nasopharyngeal/oropharyngeal swab vs Saliva | 1.655 | 1.496 | 1.83 |
| Immunocompromised status, No vs Yes | 1.067 | 0.847 | 1.343 |
| History of COVID-19 diagnosis, No vs Yes | 1.549 | 1.415 | 1.696 |

Supplementary Table 32. Full model for 3-dose VE against omicron variant infection (subgroup of Non-Hispanic and others)

| Odds Ratio Estimates | | | |
|---|---|---|---|
| Effect | Point Estimate | 95% Wald Confidence Interval | |
| Vaccination status, Vaccinated vs Unvaccinated | 0.286 | 0.262 | 0.312 |
| History of SARS-CoV-2 molecular test, No vs Yes | 0.978 | 0.905 | 1.056 |
| Preventive Care, No vs Yes | 1.067 | 0.981 | 1.161 |
| Number of outpatient and virtual visits, 0 vs >=11 | 1.519 | 1.281 | 1.802 |
| Number of outpatient and virtual visits, 1-4 vs >=11 | 1.35 | 1.218 | 1.496 |
| Number of outpatient and virtual visits, 5-10 vs >=11 | 1.187 | 1.085 | 1.299 |
| Obese, No vs Yes | 1.025 | 0.95 | 1.106 |
| Obese, Unknown vs Yes | 0.933 | 0.826 | 1.053 |
| Charlson comorbidity score, 0 vs >=2 | 1.075 | 0.933 | 1.239 |
| Charlson comorbidity score, 1 vs >=2 | 0.939 | 0.805 | 1.096 |
| Frailty, Quartile 1 vs Quartile 4 | 1.093 | 0.979 | 1.22 |
| Frailty, Quartile 2 vs Quartile 4 | 1.092 | 0.97 | 1.231 |
| Frailty, Quartile 3 vs Quartile 4 | 1.074 | 0.97 | 1.189 |
| Specimen type, Nasopharyngeal/oropharyngeal swab vs Saliva | 1.666 | 1.532 | 1.811 |
| Immunocompromised status, No vs Yes | 1.131 | 0.925 | 1.382 |
| History of COVID-19 diagnosis, No vs Yes | 1.379 | 1.245 | 1.528 |

Supplementary Table 33. Full model for 3-dose VE against omicron variant infection (subgroup of immunocompromised population)

| Odds Ratio Estimates | | | |
|---|---|---|---|
| Effect | Point Estimate | 95% Wald Confidence Interval | |
| Vaccination status, Vaccinated vs Unvaccinated | 0.706 | 0.5 | 0.997 |
| History of SARS-CoV-2 molecular test, No vs Yes | 1.271 | 0.899 | 1.798 |
| Preventive Care, No vs Yes | 1.62 | 1.088 | 2.414 |
| Number of outpatient and virtual visits, 0 vs >=11 | 0.538 | 0.097 | 2.983 |
| Number of outpatient and virtual visits, 1-4 vs >=11 | 1.174 | 0.644 | 2.141 |
| Number of outpatient and virtual visits, 5-10 vs >=11 | 1.407 | 0.967 | 2.046 |
| Obese, No vs Yes | 0.947 | 0.703 | 1.275 |
| Obese, Unknown vs Yes | 0.732 | 0.256 | 2.095 |
| Charlson comorbidity score, 0 vs >=2 | 1.03 | 0.708 | 1.501 |
| Charlson comorbidity score, 1 vs >=2 | 0.853 | 0.561 | 1.297 |
| Frailty, Quartile 1 vs Quartile 4 | 1.15 | 0.711 | 1.859 |
| Frailty, Quartile 2 vs Quartile 4 | 1.336 | 0.857 | 2.082 |
| Frailty, Quartile 3 vs Quartile 4 | 1.316 | 0.868 | 1.996 |
| Specimen type, Nasopharyngeal/oropharyngeal swab vs Saliva | 1.161 | 0.796 | 1.693 |
| History of COVID-19 diagnosis, No vs Yes | 1.413 | 0.91 | 2.193 |
| Age group, 18-44 years old vs 75 and older | 0.595 | 0.309 | 1.145 |
| Age group, 45-64 years old vs 75 and older | 0.902 | 0.482 | 1.688 |
| Age group, 65-74 years old vs 75 and older | 1.27 | 0.629 | 2.563 |
| Sex, Female vs Male | 1.033 | 0.764 | 1.396 |
| Race/ethnicity, Hispanic vs Other/Unknown | 1.122 | 0.542 | 2.319 |
| Race/ethnicity, Non-Hispanic Asian vs Other/Unknown | 1.021 | 0.422 | 2.471 |
| Race/ethnicity, Non-Hispanic Black vs Other/Unknown | 1.083 | 0.488 | 2.402 |
| Race/ethnicity, Non-Hispanic White vs Other/Unknown | 1.019 | 0.488 | 2.13 |

Supplementary Table 34. Full model for 3-dose VE against omicron variant infection (subgroup of non-immunocompromised)

| Odds Ratio Estimates | | | |
|---|---|---|---|
| **Effect** | **Point Estimate** | **95% Wald Confidence Interval** | |
| Vaccination status, Vaccinated vs Unvaccinated | 0.295 | 0.276 | 0.314 |
| History of SARS-CoV-2 molecular test, No vs Yes | 0.908 | 0.857 | 0.963 |
| Preventive Care, No vs Yes | 1.068 | 1.005 | 1.134 |
| Number of outpatient and virtual visits, 0 vs >=11 | 1.57 | 1.389 | 1.774 |
| Number of outpatient and virtual visits, 1-4 vs >=11 | 1.378 | 1.28 | 1.484 |
| Number of outpatient and virtual visits, 5-10 vs >=11 | 1.21 | 1.132 | 1.294 |
| Obese, No vs Yes | 1.015 | 0.962 | 1.071 |
| Obese, Unknown vs Yes | 0.978 | 0.897 | 1.066 |
| Charlson comorbidity score, 0 vs >=2 | 1.116 | 0.999 | 1.247 |
| Charlson comorbidity score, 1 vs >=2 | 1.03 | 0.914 | 1.16 |
| Frailty, Quartile 1 vs Quartile 4 | 1.064 | 0.984 | 1.151 |
| Frailty, Quartile 2 vs Quartile 4 | 1.063 | 0.975 | 1.16 |
| Frailty, Quartile 3 vs Quartile 4 | 1.087 | 1.01 | 1.17 |
| Specimen type, Nasopharyngeal/oropharyngeal swab vs Saliva | 1.705 | 1.6 | 1.817 |
| History of COVID-19 diagnosis, No vs Yes | 1.478 | 1.382 | 1.581 |
| Age group, 18-44 years old vs 75 and older | 0.37 | 0.307 | 0.445 |
| Age group, 45-64 years old vs 75 and older | 0.526 | 0.439 | 0.632 |
| Age group, 65-74 years old vs 75 and older | 0.832 | 0.685 | 1.009 |
| Sex, Female vs Male | 1.141 | 1.084 | 1.2 |
| Race/ethnicity, Hispanic vs Other/Unknown | 0.968 | 0.882 | 1.063 |
| Race/ethnicity, Non-Hispanic Asian vs Other/Unknown | 1.472 | 1.297 | 1.67 |
| Race/ethnicity, Non-Hispanic Black vs Other/Unknown | 0.931 | 0.83 | 1.044 |
| Race/ethnicity, Non-Hispanic White vs Other/Unknown | 1.015 | 0.921 | 1.119 |

Supplementary Table 35. Full model for 3-dose VE against omicron variant infection among individuals without COVID-19 history, age<65

**Odds Ratio Estimates**

| Effect | Point Estimate | 95% Wald Confidence Interval | |
|---|---|---|---|
| Vaccination status, Vaccinated vs Unvaccinated | 0.299 | 0.279 | 0.32 |
| History of SARS-CoV-2 molecular test, No vs Yes | 0.898 | 0.844 | 0.955 |
| Preventive Care, No vs Yes | 1.087 | 1.016 | 1.163 |
| Number of outpatient and virtual visits, 0 vs >=11 | 1.577 | 1.373 | 1.81 |
| Number of outpatient and virtual visits, 1-4 vs >=11 | 1.439 | 1.323 | 1.566 |
| Number of outpatient and virtual visits, 5-10 vs >=11 | 1.28 | 1.185 | 1.382 |
| Obese, No vs Yes | 1.026 | 0.965 | 1.092 |
| Obese, Unknown vs Yes | 0.974 | 0.885 | 1.072 |
| Charlson comorbidity score, 0 vs >=2 | 1.151 | 1.003 | 1.322 |
| Charlson comorbidity score, 1 vs >=2 | 1.049 | 0.901 | 1.22 |
| Frailty, Quartile 1 vs Quartile 4 | 1.032 | 0.946 | 1.126 |
| Frailty, Quartile 2 vs Quartile 4 | 1.062 | 0.962 | 1.172 |
| Frailty, Quartile 3 vs Quartile 4 | 1.052 | 0.968 | 1.142 |
| Specimen type, Nasopharyngeal/oropharyngeal swab vs Saliva | 1.727 | 1.607 | 1.855 |
| Immunocompromised status, No vs Yes | 1.035 | 0.865 | 1.239 |
| Age Group, 18-44 years old vs 45-64 years old | 0.697 | 0.653 | 0.744 |
| Sex, Female vs Male | 1.144 | 1.079 | 1.212 |
| Race/ethnicity, Hispanic vs Other/Unknown | 0.98 | 0.885 | 1.085 |
| Race/ethnicity, Non-Hispanic Asian vs Other/Unknown | 1.475 | 1.285 | 1.694 |
| Race/ethnicity, Non-Hispanic Black vs Other/Unknown | 0.927 | 0.816 | 1.052 |
| Race/ethnicity, Non-Hispanic White vs Other/Unknown | 1.027 | 0.923 | 1.143 |

Supplementary Table 36. Full model for 3-dose VE against omicron variant infection among individuals without COVID-19 history, age≥65

| Odds Ratio Estimates | | | |
|---|---|---|---|
| Effect | Point Estimate | 95% Wald Confidence Interval | |
| Vaccination status, Vaccinated vs Unvaccinated | 0.355 | 0.279 | 0.451 |
| History of SARS-CoV-2 molecular test, No vs Yes | 1.14 | 0.95 | 1.368 |
| Preventive Care, No vs Yes | 0.791 | 0.564 | 1.109 |
| Number of outpatient and virtual visits, 0 vs >=11 | 0.774 | 0.326 | 1.842 |
| Number of outpatient and virtual visits, 1-4 vs >=11 | 1.034 | 0.738 | 1.449 |
| Number of outpatient and virtual visits, 5-10 vs >=11 | 0.864 | 0.699 | 1.069 |
| Obese, No vs Yes | 0.836 | 0.695 | 1.006 |
| Obese, Unknown vs Yes | 0.715 | 0.417 | 1.226 |
| Charlson comorbidity score, 0 vs >=2 | 1.182 | 0.933 | 1.497 |
| Charlson comorbidity score, 1 vs >=2 | 1.001 | 0.794 | 1.262 |
| Frailty, Quartile 1 vs Quartile 4 | 1.009 | 0.748 | 1.362 |
| Frailty, Quartile 2 vs Quartile 4 | 1.082 | 0.833 | 1.405 |
| Frailty, Quartile 3 vs Quartile 4 | 1.029 | 0.806 | 1.316 |
| Specimen type, Nasopharyngeal/oropharyngeal swab vs Saliva | 1.33 | 1.061 | 1.669 |
| Immunocompromised status, No vs Yes | 1.149 | 0.802 | 1.646 |
| Age group, 65-74 years old vs 75 and older | 0.846 | 0.694 | 1.031 |
| Sex, Female vs Male | 0.945 | 0.795 | 1.124 |
| Race/ethnicity, Hispanic vs Other/Unknown | 0.996 | 0.588 | 1.688 |
| Race/ethnicity, Non-Hispanic Asian vs Other/Unknown | 1.037 | 0.59 | 1.823 |
| Race/ethnicity, Non-Hispanic Black vs Other/Unknown | 0.899 | 0.511 | 1.58 |
| Race/ethnicity, Non-Hispanic White vs Other/Unknown | 0.952 | 0.566 | 1.6 |
